# District Level Variation in Hypertension Epidemiology in India and Influence of Social Determinants: National Family Health Survey-5

**DOI:** 10.1101/2023.10.02.23296421

**Authors:** Rajeev Gupta, Kiran Gaur, Suresh C Sharma, Raghubir S Khedar, Rajinder K Dhamija

## Abstract

**BACKGROUND:** Enumeration of state and district-level variation in hypertension prevalence in India and to evaluate the influence of social determinants.

**METHODS:** We used data from the Fifth National Family Health Survey (NFHS-5) from 707 districts and 825,954 participants (women 724,115, men 101,839 men) on prevalence of hypertension defined according to standard criteria. Data on multiple social determinants were also obtained from NFHS-5 report.

**RESULTS:** Age-standardized prevalence of hypertension was 22.4% (women 21.3%, men 24.0%) with the highest prevalence in women and men, respectively, in Sikkim (34.5 and 41.6%) and Punjab (31.2 and 37.7%) and lowest in Rajasthan (15.4 and 17.9%) and Ladakh (15.7 and 17.4%). Prevalence was more in western and southern Indian districts. High prevalence of hypertension in the young (<30y) was observed in northeastern and northern states. District-level hypertension prevalence correlated negatively with multi-dimensional poverty index (R^2^ women 0.299, men 0.245) and positively with female literacy (women 0.165, men 0.134). Among women, districts with the highest availability of electricity, clean water, sanitation, clean cooking fuels, healthcare service delivery and better nutrition were associated with more hypertension on univariate and multivariate analyses (p<0.05).

**CONCLUSIONS:** The study shows significant geographical variation in hypertension prevalence in India. Hypertension is more in men with high prevalence of premature hypertension. Better district-level development (less poverty, more literacy) and healthcare services are associated with greater hypertension prevalence in women.

**SUMMARY TABLE:** *What is known about the topic:*

- Significant state-level variation in hypertension prevalence in India has been reported but district-level variation is not known.
- Social determinants are important in hypertension but not well studied, especially in women.

*What this study adds:*

- The study shows a significant district-level variation with greater hypertension prevalence in southern and western India.
- Hypertension among the young, <30 years, is more in less developed districts.
- Social determinants of hypertension in women are less poverty, more literacy and availability of healthcare services.

## INTRODUCTION

World Health Organization (WHO) and others have reported that high blood pressure (BP) is the most important risk factor for mortality and disease burden in both women and men, globally and in India.^1-4^ Studies have reported significant inter-country variation in hypertension prevalence with the highest prevalence rates in Eastern European, Central Asian and Eastern Asian countries.^1,3,5,6^ Within-country variation has also been reported from several large countries-China, USA, Brazil, Indonesia, Pakistan, UK- and Europe.^7-13^ A county (district) level variation has been reported from the USA and UK.^14,15^ Studies from USA, UK and China have reported greater hypertension prevalence in less developed regions and counties.^7,8,12^ Risk factors for hypertension are social and biological.^16^ Social determinants of hypertension include low-quality urban and rural infrastructure, low socioeconomic status, social disorganization, unemployment, adverse work environment, illiteracy, adverse early life events, subnormal maternal and child health, etc.^16,17^ It has been reported that macrolevel and micro-level variations in social, lifestyle and biological factors are responsible for difference in hypertension prevalence.^16^

Previous national studies in India-District Level Health Survey/Adult Health Survey (DLHS-4/AHS-3)^18^ and Fourth National Family Health Survey (NFHS-4)^19^ have reported significant state-level variation in hypertension prevalence. It has been reported that macro-level social factors such as greater urbanization, better human development index, more literacy and raised body mass index (BMI) are associated with this variation.^19^ This contrasts with developed countries where hypertension is more in lesser developed and deprived regions and communities.^12,15^ India has undergone rapid socio-economic progress in recent years. To correlate variation in hypertension prevalence at the district (county) level with various social factors we performed the present study. We used district-level data from Fifth National Family Health Survey (NFHS-5)^20^ on hypertension prevalence and performed correlation with multiple social determinants of health.

## METHODS

Data from NFHS-5 are available at the National Family Health Survey website at http://rchiips.org/nfhs/factsheet_NFHS-5.shtml.^20^ NFHS is a large-scale, multi-round survey conducted in a representative sample of households throughout India. Multiple rounds of the survey have been conducted since the first survey in 1992-93. The survey provides state and national information for India on fertility, infant and child mortality, the practice of family planning, maternal and child health, reproductive health, nutrition, anemia, utilization and quality of health and family planning services. Each successive round of the NFHS has had two specific goals: a) to provide essential data on health and family welfare needed by the Ministry of Health and Family Welfare and other agencies for policy and program purposes, and b) to provide information on important emerging health and family welfare issues. Ethics clearance has been obtained by the Institutional Ethics Committee of Indian Institute of Population Sciences, Mumbai, India. Technical assistance for the NFHS has been provided by ICF International (Virginia, USA) and other organizations on specific issues. The funding for different rounds of NFHS has been provided by USAID, DFID, the Bill and Melinda Gates Foundation, UNICEF, UNFPA, and Ministry of Health and Family Welfare, Government of India. In NFHS-5, similar to the previous iterations, the sample was designed to provide data of all key indicators at the national and state levels as well as estimates for most key indicators at the district level for almost all the districts of the country.^20^

The total sample size in NFHS-5 was 610,000 households in India. This was estimated to produce reliable data for each district of the country. The sample selection process was similar to the previous NFHS studies. The rural sample was selected through a 2-stage sampling with villages as the primary sampling unit (PSU) at the first stage selected with probability proportional to size, followed by a random selection of 22 households in each PSU at the second stage. In urban areas, there was also a 2-stage sampling design with Census Enumeration Blocks (CEB) selected at the first stage and a random selection of 22 households in each CEB at the second stage. In the second stage in both rural and urban areas, households were selected after conducting a complete mapping and household listing operation in the selected first-stage units. NFHS-5 fieldwork for India was conducted in two phases, phase one from 17 June 2019 to 30 January 2020 and phase two from 2 January 2020 to 30 April 2021 by 17 Field Agencies and gathered information from 636,699 households, 724,115 women, and 101,839 men in 705 districts of the country.

The biomarker questionnaire included measurements of height, weight, waist and hip circumference, hemoglobin levels and finger-stick blood for additional testing in a laboratory for women aged 15-49 years and men aged 15-54 years. Blood pressure and random blood glucose were measured in all women and men aged 15 years and more in contrast to NFHS-4.^19^ Questionnaire information and biomarkers were collected after informed consent from each participant. In the NFHS-5 the BP of eligible respondents was measured using an OMRON Blood Pressure Monitor to determine the prevalence of hypertension. BP measurements for each respondent were taken on three separate occasions and the readings were recorded in the Biomarker Questionnaire with an interval of 5 minutes between readings. Respondents whose systolic BP was >130 mm Hg or diastolic BP > 85 mm Hg were considered to have elevated blood pressure readings and were encouraged to see a doctor for a full evaluation. Hypertension was diagnosed when systolic BP was ≥140 mmHg and/or diastolic BP ≥90 mmHg or when the participant was taking medicine to control BP and reported in per cent (%).

### Statistical analyses

Data sheets were downloaded from the NFHS-5 website. Additional data were obtained from the Population Research Centre, Institute of Economic Growth, Delhi University, New Delhi, India. Data were transferred to MS Excel sheets and descriptive analyses were performed using this program. Hypertension prevalence rates are reported separately for women and men and have been age-adjusted to the population of India. A χ^2^ test was used to determine the significance of women-men differences. Hypertension prevalence rates for various districts of the country (n=705) were also obtained from the NFHS-5 website and reported separately for women and men. State-level mean, medians and 25-75 interquartile intervals (IQR) for hypertension were calculated. Univariate correlation of district hypertension prevalence with social determinants (Supplementary Table 1) was performed using polynomial regression and graphs were computed in MS Excel. For multivariate correlation, we used SPSS statistical package.

## RESULTS

NFHS-5 enrolled 724,115 women and 101,839 men >15 years of age (total of 825,954) with response rates more than 90%.^20^ Data for multiple socioeconomic, lifestyle and clinical variables were obtained and recorded. National age-adjusted prevalence (%) of hypertension among women was 21.3% and men 24.0% (overall 22.4%). Age-adjusted prevalence (%) of hypertension among women and men in different states of India is in Table 1. There is significant state-level variation in hypertension prevalence. In women the highest prevalence is in Sikkim (34.5%), Punjab (31.2%) and Goa (27.5%) and the lowest in Rajasthan (15.4%), Ladakh (15.7%) and Bihar (15.9%). In men the highest prevalence is in Sikkim (41.6%), Punjab (37.7%) and Manipur (33.2%) while the lowest is in Ladakh (17.4%), Rajasthan (17.9%) and Bihar (18.4%) (Figure 1). Hypertension prevalence in various states and districts of India in women and men is shown as a heat map (Figure 2). High prevalence is observed in southern and eastern Indian states. There is a significant district-level variation with greater prevalence in most western and southern districts in women and western, southern and eastern districts in men. Absolute prevalence rates for each district among women and men are in Supplementary Tables 2-34. We also calculated median (IQR) hypertension prevalence in each state according to the district-level prevalence and prevalence rates for women and men shown in Supplementary Figure 1.

**Table 1:** Hypertension prevalence in Indian states in NFHS-5 in women and men ≥15 years. Criteria for diagnosis of hypertension were systolic BP ≥140 mmHg and/or diastolic BP ≥90 mmHg or on medical treatment.

|  | Sample size |  | Age-standardized hypertension prevalence (%) |  |  |
| --- | --- | --- | --- | --- | --- |
|  | Women | Men | Women | Men | Combined |
| **India** | **624115** | **101839** | **21.3** | **24.0** | **22.4** |
| Andaman Nicobar Islands | 2397 | 367 | 25.3 | 30.2 | 26.0 |
| Andhra Pradesh | 10975 | 1558 | 25.3 | 29 | 25.8 |
| Arunachal Pradesh | 19765 | 2881 | 24.9 | 33.1 | 25.9 |
| Assam | 34979 | 4973 | 19.1 | 20.3 | 19.2 |
| Bihar | 42483 | 4897 | 15.9 | 18.4 | 16.2 |
| Chandigarh | 746 | 104 | 25 | 30.6 | 25.7 |
| Chhattisgarh | 28468 | 4174 | 23.6 | 27.7 | 24.1 |
| Delhi | 11159 | 1700 | 24.1 | 32.8 | 25.3 |
| Goa | 2030 | 313 | 27.5 | 26.8 | 27.4 |
| Gujarat | 33343 | 5351 | 20.6 | 20.3 | 20.6 |
| Haryana | 21909 | 3224 | 21 | 25.1 | 21.5 |
| Himachal Pradesh | 10368 | 1477 | 22.2 | 24.4 | 22.5 |
| Jammu & Kashmir | 23037 | 3087 | 20 | 18.9 | 19.9 |
| Jharkhand | 26495 | 3414 | 17.8 | 22.6 | 18.3 |
| Karnataka | 30455 | 4516 | 25 | 26.9 | 25.2 |
| Kerala | 10969 | 1473 | 30.9 | 32.8 | 31.1 |
| Lakshadweep | 1234 | 135 | 24.8 | 24.7 | 24.8 |
| Madhya Pradesh | 48410 | 7025 | 20.6 | 22.7 | 20.9 |
| Maharashtra | 33755 | 5497 | 23.1 | 24.4 | 23.3 |
| Manipur | 8042 | 1162 | 23 | 33.2 | 24.3 |
| Meghalaya | 13089 | 1824 | 18.7 | 21.4 | 19.0 |
| Mizoram | 7279 | 1105 | 17.7 | 25.2 | 18.7 |
| Nagaland | 9694 | 1456 | 22.4 | 28.7 | 23.2 |
| Odisha | 27971 | 3865 | 22.4 | 25.6 | 22.8 |
| Puducherry | 3669 | 534 | 23 | 30.1 | 23.9 |
| Punjab | 21771 | 3296 | 31.2 | 37.7 | 32.1 |
| Rajasthan | 42990 | 6353 | 15.4 | 17.9 | 15.7 |
| Sikkim | 3271 | 469 | 34.5 | 41.6 | 35.4 |
| Tamil Nadu | 25650 | 3372 | 24.8 | 30.2 | 25.4 |
| Telangana | 27518 | 3863 | 26.1 | 31.4 | 26.8 |
| Tripura | 7314 | 990 | 20.9 | 22.7 | 21.1 |
| Uttarakhand | 13280 | 1586 | 22.9 | 31.8 | 23.8 |
| Uttar Pradesh | 93124 | 12043 | 18.4 | 21.7 | 18.8 |
| West Bengal | 21408 | 3021 | 20.5 | 20.1 | 20.5 |

**Figure 1:** Hypertension prevalence among women and men in various states of India in NFHS-5.

**Figure 2:** Heat map showing state-level prevalence (Figure 2a) and district-level prevalence (Figure 2b) of hypertension among women and men in NFHS-5.

Age-group stratified hypertension prevalence among women and men for the whole country and all the states are in Table 2. There is a significant age-associated escalation in its prevalence in all the states of the country (Mantel-Haenszel χ^2^ for trend, p <0.001). Higher prevalence of hypertension in the young (<30 years) is observed among men and women in most North-Eastern Indian states (Arunachal Pradesh, Assam, Meghalaya, Nagaland, Sikkim and West Bengal), Jammu & Kashmir, Ladakh and Goa (Supplementary Figure 2). On the other hand, high prevalence of hypertension among the elderly (≥60yr) is observed in Delhi, Goa, Karnataka, Kerala, Punjab, Sikkim and Telangana.

**Table 2:**
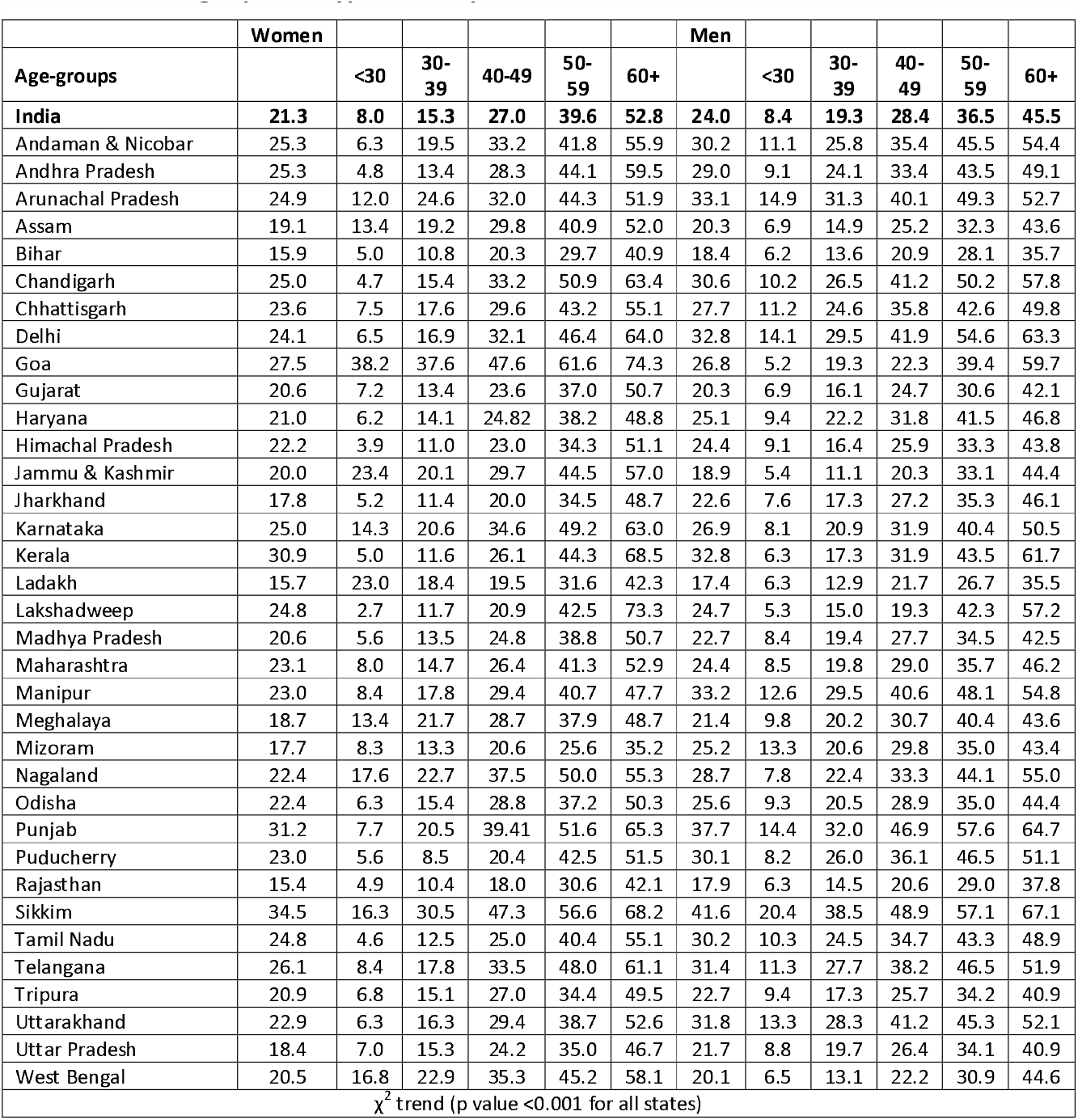
Age-group specific hypertension prevalence in Indian states in women and men.

|  | Women |  |  |  |  |  | Men |  |  |  |  |  |
| --- | --- | --- | --- | --- | --- | --- | --- | --- | --- | --- | --- | --- |
| Age-groups |  | <30 | 30-39 | 40-49 | 50-59 | 60+ |  | <30 | 30-39 | 40-49 | 50-59 | 60+ |
| **India** | **21.3** | **8.0** | **15.3** | **27.0** | **39.6** | **52.8** | **24.0** | **8.4** | **19.3** | **28.4** | **36.5** | **45.5** |
| Andaman & Nicobar | 25.3 | 6.3 | 19.5 | 33.2 | 41.8 | 55.9 | 30.2 | 11.1 | 25.8 | 35.4 | 45.5 | 54.4 |
| Andhra Pradesh | 25.3 | 4.8 | 13.4 | 28.3 | 44.1 | 59.5 | 29.0 | 9.1 | 24.1 | 33.4 | 43.5 | 49.1 |
| Arunachal Pradesh | 24.9 | 12.0 | 24.6 | 32.0 | 44.3 | 51.9 | 33.1 | 14.9 | 31.3 | 40.1 | 49.3 | 52.7 |
| Assam | 19.1 | 13.4 | 19.2 | 29.8 | 40.9 | 52.0 | 20.3 | 6.9 | 14.9 | 25.2 | 32.3 | 43.6 |
| Bihar | 15.9 | 5.0 | 10.8 | 20.3 | 29.7 | 40.9 | 18.4 | 6.2 | 13.6 | 20.9 | 28.1 | 35.7 |
| Chandigarh | 25.0 | 4.7 | 15.4 | 33.2 | 50.9 | 63.4 | 30.6 | 10.2 | 26.5 | 41.2 | 50.2 | 57.8 |
| Chhattisgarh | 23.6 | 7.5 | 17.6 | 29.6 | 43.2 | 55.1 | 27.7 | 11.2 | 24.6 | 35.8 | 42.6 | 49.8 |
| Delhi | 24.1 | 6.5 | 16.9 | 32.1 | 46.4 | 64.0 | 32.8 | 14.1 | 29.5 | 41.9 | 54.6 | 63.3 |
| Goa | 27.5 | 38.2 | 37.6 | 47.6 | 61.6 | 74.3 | 26.8 | 5.2 | 19.3 | 22.3 | 39.4 | 59.7 |
| Gujarat | 20.6 | 7.2 | 13.4 | 23.6 | 37.0 | 50.7 | 20.3 | 6.9 | 16.1 | 24.7 | 30.6 | 42.1 |
| Haryana | 21.0 | 6.2 | 14.1 | 24.82 | 38.2 | 48.8 | 25.1 | 9.4 | 22.2 | 31.8 | 41.5 | 46.8 |
| Himachal Pradesh | 22.2 | 3.9 | 11.0 | 23.0 | 34.3 | 51.1 | 24.4 | 9.1 | 16.4 | 25.9 | 33.3 | 43.8 |
| Jammu & Kashmir | 20.0 | 23.4 | 20.1 | 29.7 | 44.5 | 57.0 | 18.9 | 5.4 | 11.1 | 20.3 | 33.1 | 44.4 |
| Jharkhand | 17.8 | 5.2 | 11.4 | 20.0 | 34.5 | 48.7 | 22.6 | 7.6 | 17.3 | 27.2 | 35.3 | 46.1 |
| Karnataka | 25.0 | 14.3 | 20.6 | 34.6 | 49.2 | 63.0 | 26.9 | 8.1 | 20.9 | 31.9 | 40.4 | 50.5 |
| Kerala | 30.9 | 5.0 | 11.6 | 26.1 | 44.3 | 68.5 | 32.8 | 6.3 | 17.3 | 31.9 | 43.5 | 61.7 |
| Ladakh | 15.7 | 23.0 | 18.4 | 19.5 | 31.6 | 42.3 | 17.4 | 6.3 | 12.9 | 21.7 | 26.7 | 35.5 |
| Lakshadweep | 24.8 | 2.7 | 11.7 | 20.9 | 42.5 | 73.3 | 24.7 | 5.3 | 15.0 | 19.3 | 42.3 | 57.2 |
| Madhya Pradesh | 20.6 | 5.6 | 13.5 | 24.8 | 38.8 | 50.7 | 22.7 | 8.4 | 19.4 | 27.7 | 34.5 | 42.5 |
| Maharashtra | 23.1 | 8.0 | 14.7 | 26.4 | 41.3 | 52.9 | 24.4 | 8.5 | 19.8 | 29.0 | 35.7 | 46.2 |
| Manipur | 23.0 | 8.4 | 17.8 | 29.4 | 40.7 | 47.7 | 33.2 | 12.6 | 29.5 | 40.6 | 48.1 | 54.8 |
| Meghalaya | 18.7 | 13.4 | 21.7 | 28.7 | 37.9 | 48.7 | 21.4 | 9.8 | 20.2 | 30.7 | 40.4 | 43.6 |
| Mizoram | 17.7 | 8.3 | 13.3 | 20.6 | 25.6 | 35.2 | 25.2 | 13.3 | 20.6 | 29.8 | 35.0 | 43.4 |
| Nagaland | 22.4 | 17.6 | 22.7 | 37.5 | 50.0 | 55.3 | 28.7 | 7.8 | 22.4 | 33.3 | 44.1 | 55.0 |
| Odisha | 22.4 | 6.3 | 15.4 | 28.8 | 37.2 | 50.3 | 25.6 | 9.3 | 20.5 | 28.9 | 35.0 | 44.4 |
| Punjab | 31.2 | 7.7 | 20.5 | 39.41 | 51.6 | 65.3 | 37.7 | 14.4 | 32.0 | 46.9 | 57.6 | 64.7 |
| Puducherry | 23.0 | 5.6 | 8.5 | 20.4 | 42.5 | 51.5 | 30.1 | 8.2 | 26.0 | 36.1 | 46.5 | 51.1 |
| Rajasthan | 15.4 | 4.9 | 10.4 | 18.0 | 30.6 | 42.1 | 17.9 | 6.3 | 14.5 | 20.6 | 29.0 | 37.8 |
| Sikkim | 34.5 | 16.3 | 30.5 | 47.3 | 56.6 | 68.2 | 41.6 | 20.4 | 38.5 | 48.9 | 57.1 | 67.1 |
| Tamil Nadu | 24.8 | 4.6 | 12.5 | 25.0 | 40.4 | 55.1 | 30.2 | 10.3 | 24.5 | 34.7 | 43.3 | 48.9 |
| Telangana | 26.1 | 8.4 | 17.8 | 33.5 | 48.0 | 61.1 | 31.4 | 11.3 | 27.7 | 38.2 | 46.5 | 51.9 |
| Tripura | 20.9 | 6.8 | 15.1 | 27.0 | 34.4 | 49.5 | 22.7 | 9.4 | 17.3 | 25.7 | 34.2 | 40.9 |
| Uttarakhand | 22.9 | 6.3 | 16.3 | 29.4 | 38.7 | 52.6 | 31.8 | 13.3 | 28.3 | 41.2 | 45.3 | 52.1 |
| Uttar Pradesh | 18.4 | 7.0 | 15.3 | 24.2 | 35.0 | 46.7 | 21.7 | 8.8 | 19.7 | 26.4 | 34.1 | 40.9 |
| West Bengal | 20.5 | 16.8 | 22.9 | 35.3 | 45.2 | 58.1 | 20.1 | 6.5 | 13.1 | 22.2 | 30.9 | 44.6 |
| $\chi^2$ trend (p value <0.001 for all states) | | | | | | | | | | | | |

Data on multiple social determinants were obtained from various government databases, NFHS-5 and others.^19,20^ District-level multidimensional poverty index is negatively associated with the prevalence of hypertension in both women and men with polynomial R^2^ of 0.299 and 0.245, respectively (Figure 3a). Hypertension prevalence is positively associated with district-level female literacy levels in both women and men with polynomial R^2^ of 0.165 and 0.134, respectively (Figure 3b). Data on social determinants in NFHS-5 have been primarily obtained for women, therefore, we determined variation of hypertension prevalence in women according to them. Correlation of district-level hypertension prevalence among women with various social, healthcare delivery and clinical determinants are in Figure 4. Increasing hypertension prevalence among women correlates significantly with declining poverty, better literacy, sanitation, and electricity, and availability of clean fuel for cooking (Figure 4a). Greater hypertension prevalence is also observed in district with lower childhood marriages and teenage pregnancies and greater vaccination coverage, modern contraceptive usage and availability of skilled delivery services (Figure 4b). Presence of childhood overweight, adult overweight (BMI ≥25) and adult hyperglycemia is also associated with greater hypertension prevalence (Figure 4c).

**Figure 3:** Inverse correlation of district-level hypertension prevalence in women and men with increasing multidimensional poverty index (Figure 3a) and positive correlation of district-level hypertension prevalence in women and men with increasing literacy (Figure 3b).

**Figure 4.** (a-c): Correlation of district-level hypertension prevalence in women with various (a) social, (b) healthcare delivery, and (c) clinical determinants.

To identify important social determinants among women we classified various socioeconomic-, environmental-, healthcare service-, womankind specific-, and nutritional variables into tertiles and determined hypertension prevalence in each of them (Table 3). Analyses of social determinants show that in districts with lower poverty and illiteracy and the highest prevalence of availability of electricity, clean water, sanitation and clean cooking fuels, the prevalence of hypertension is the highest. Availability of better healthcare service delivery (higher availability of skilled childbirth, use of modern contraception and full vaccination) and better nutrition (lower anemia, childhood stunting or wasting, overweight in children and adults) is also associated with greater prevalence of hypertension. We also calculated univariate and multivariate regression coefficients to identify important variables (Table 4). On univariate analysis. There are multiple factors associated with district level hypertension prevalence including socioeconomic and environmental variables (multidimensional poverty index, illiteracy, access to electricity, clean drinking water and sanitation, tobacco use), healthcare service delivery parameters and nutritional factors. However, on multivariate analyses risk factors that continue to be significant are low female illiteracy (standardized b -0.12), improved drinking water (0.11), sanitation (0.19), modern contraceptive use (0.20) and overweight or obesity in women (0.44) (p<0.05).

**Table 3:** Hypertension prevalence (%, 95% CI) in women according to tertiles of various social indicators.

| Variable | Hypertension prevalence (%) |  |  | F-ratio (p value) |
| --- | --- | --- | --- | --- |
|  | Low tertile | Middle tertile | High tertile |  |
| Socioeconomic and environmental determinants |  |  |  |  |
| Multidimensional poverty index | 22.8(17.7-28.0) | 18.6(14.6-22.6) | 17.7(14.2-21.1) | 66.72 (0.00) |
| Illiteracy (women) | 18.4(14.5-22.4) | 20.4(15.8-25.1) | 23.8(18.3-29.2) | 52.77(0.00) |
| Health insurance status (any member) | 21.2(15.7-26.6) | 22.0(17.1-26.9) | 20.7(15.6-25.8) | 2.95(0.05) |
| Electricity access | 18.8(14.5-23.2) | 18.6(14.0-23.3) | 21.6(16.4-26.8) | 6.83(0.00) |
| Improved drinking water | 18.0(15.1-21.0) | 20.4(15.9-26.8) | 24.8(21.0-26.8) | 2.70(0.06) |
| Improved sanitation | 167.4(14.0-20.9) | 20.1(16.0-24.1) | 23.8(18.1-29.4) | 76.36(0.00) |
| Clean fuel for cooking | 19.2(15.1-23.3) | 20.7(15.5-25.9) | 24.5(19.6-29.4) | 78.03(0.00) |
| Tobacco use (women) | 21.5(16.2-26.8) | 21.2(17.4-25.0) | 19.0(13.8-24.1) | 2.77(0.06) |
| Healthcare services and women-specific factors |  |  |  |  |
| Skilled childbirth services | 21.0(15.7-26.3) | 18.4(14.5-22.3) | 21.6(16.4-26.9) | 9.3(0.00) |
| Modern contraception use | 19.2(15.5-23.0) | 20.7(15.3-26.0) | 22.5(17.5-27.6) | 15.55(0.00) |
| Full vaccination | 20.3(15.6-25.0) | 20.5(25.4-25.6) | 22.3(17.2-27.4) | 11.00(0.00) |
| Child marriage | 22.7(17.2-28.2) | 20.6(16.0-25.1) | 17.8(14.2-21.3) | 35.16(0.00) |
| Teenage pregnancy | 21.716.3-27.1) | 20.4(16.0-24.8) | 19.0(15.3-22.8) | 6.09(0.00) |
| Anemia (adult women) | 24.5(18.1-31.1) | 21.4(16.4-26.4) | 19.0(15.1-22.8) | 27.32(0.00) |
| Nutritional and other factors |  |  |  |  |
| Childhood stunting | 23.8(18.3-29.3) | 20.6(15.8-25.3) | 18.3(14.6-22.0) | 46.83(0.00) |
| Childhood wasting | 22.2(16.8-27.7) | 20.3(15.6-24.9) | 20.5(15.8-25.2) | 12.92(0.00) |
| Underweight children <5 years | 23.6(18.1-29.1) | 20.3(15.8-24.8) | 18.8(14.1-23.4) | 43.71(0.00) |
| Overweight children <5 years | 21.4(16.2-26.5) | 21.0(16.2-25.9) | 26.1(17.3-34.8) | 4.28(0.014) |
| Overweight adult (women) | 18.9(14.8-23.0) | 22.2(17.6-26.9) | 27.0(22.2-31.9) | 137.38(0.00) |
| Hyperglycemia (random glucose ≥140 mg/dl) | 20.1(15.4-24.7) | 23.3(18.1-28.4) | 29.7(25.2-34.1) | 63.93(0.00) |
| CI confidence intervals, |  |  |  |  |

**Table 4:** Univariate and multivariate regression analysis (standardized b) of district-level hypertension prevalence (women) with various socioeconomic and other factors.

| Variable | Univariate standardized beta coefficient (p-value) | Multivariate adjusted standardized beta coefficient (for all variables) (p value) |
| --- | --- | --- |
| **Socioeconomic and environmental determinants** |  |  |
| Multidimensional poverty index | -0.21(0.00) | -0.04(0.73) |
| Illiteracy (Women) | -0.17(0.00) | -0.12(0.00) |
| Health insurance (any member) | 0.00(0.99) | 0.01(0.90) |
| Electricity access | 0.34(0.00) | -0.04(0.44) |
| Improved drinking water | 0.06 (0.01) | 0.11(0.01) |
| Improved sanitation | 0.17(0.00) | 0.19(0.00) |
| Clean fuel for cooking | 0.10(0.00) | -0.04(0.52) |
| Tobacco use (women) | 0.05(0.00) | 0.07(0.15) |
| **Healthcare service delivery and female specific parameters** |  |  |
| Skilled delivery | 0.15(0.00) | -0.09(0.10) |
| Modern contraception use | 0.09(0.00) | 0.20(0.00) |
| Full vaccination | 0.08(0.00) | 0.04(0.38) |
| Child marriage | -0.14(0.00) | -0.12(0.09) |
| Teenage pregnancy | -0.15(0.00) | 0.06(0.33) |
| Anemia (women) | -0.12(0.00) | -0.11(0.07) |
| **Nutritional factors (female)** |  |  |
| Childhood stunting | -0.26(0.00) | -0.13(0.07) |
| Childhood wasting | -0.18(0.00) | -0.09(0.22) |
| Underweight children <5 years | -0.20(0.00) | 0.21(0.06) |
| Overweight children <5 years | 0.18(0.00) | 0.04(0.40) |
| Overweight adult women | 0.28(0.00) | 0.44(0.00) |
| Hyperglycemia women | 0.47(0.00) | 0.10(0.06) |

## DISCUSSION

Fifth National Family Health Survey (NFHS-5) is the most representative and comprehensive hypertension epidemiology study in India. The study shows that there is substantial variation in hypertension prevalence across states and districts of the country. The prevalence of hypertension is more in better-developed states and districts. Hypertension in the young (<30 y) is more in the northeastern and northern states of the country. Greater hypertension prevalence in more developed districts (lower multidimensional poverty, and better literacy, healthcare services and nutritional status) in India, contrasts with developed countries where hypertension is more in lesser developed locations and states. These findings suggest that macrolevel cardiovascular disease risk factor transition associated with demographic and epidemiological shifts is still evolving in India.

Geographic variation in hypertension prevalence has been reported from all the large countries of the world.^7-15^ There are significant difference in prevalence rates at state and district-levels. In China, greater hypertension prevalence has been reported from less developed western and south-eastern regions;^7^ in USA, hypertension prevalence is greater in southern and south-eastern states and less developed counties;^8^ and in the UK hypertension is more in deprived counties of northern region.^12^ Global Burden of Disease (GBD) Study sub-national collaborators have reported greater hypertension related mortality in deprived regions of Brazil.^9^ On the other hand, in lower middle income countries such as India, Indonesia and Pakistan, greater hypertension prevalence and hypertension related cardiovascular mortality have been reported from better developed states and locations.^10,11,19^ Country-level differences also exist in hypertension In Europe and Asia. In Europe hypertension prevalence is significantly greater in less developed countries of Central and Eastern Europe,^13^ while in Asia the prevalence of hypertension is significantly lower in high income countries of Eastern Asia and high in Central and Southern Asian countries.^21^ Positive association of hypertension in India with better development suggests that the risk factor transition that happens with social development has not yet occurred in India.^23^ There is, on the other hand, evidence that at an individual level, hypertension may be more among illiterate rural and slum-dwelling populations,^23,24^ and there is a significant association of low socioeconomic and educational status with greater cardiovascular mortality.^25,26^ In the present study we have shown a positive association of human development with hypertension in India and more studies are needed to identify the importance of individual and macrolevel social determinants.

An important finding in the present study is a high prevalence of premature hypertension, especially in lesser developed states of northeastern and northern India. Higher prevalence of hypertension at young age in India, compared to countries of North and South America and Europe has been previously reported in the DLHS-4/AHS study.^18^ Northeastern Indian states have a very proportion of tribal population and a high prevalence of hypertension among these populations has also been reported.^27^ Hypertension in the young is an important problem among less developed societies in Africa and among African Americans in USA.^28^ There is a significant association of premature hypertension with stroke in these communities. Million Death Study in India reported a significantly greater stroke burden and mortality in the eastern and north-eastern states of the country.^29^ This corresponds to the presence of premature hypertension in these regions. More studies are required to elucidate reasons for greater hypertension and stroke burden among the Sub-Himalayan regions of India.

The study has several strengths-this is the one of the first studies that has evaluated hypertension prevalence in each district of the country. The DLHS-4/AHS study also evaluated hypertension in multiple districts but two important states were missing.^18^ IN NFHS-4 the sample size was restricted to the young and middle aged and did not provide data on elderly which have greater hypertension.^30^ The ICMR-INDIAB study also obtained data on hypertension in multiple states but sampling was based on state-level data and not at the district level.^31^ We have also identified districts with greater hypertension prevalence. This is important for policy-making as these districts should be the initial focus for hypertension control initiatives.^32^ Identification of districts with premature hypertension is also important as deployment of primary prevention strategies in these regions (salt, alcohol and tobacco control, healthy foods). Limitations of the study include a skewed female-male ratio in the recruitment of the sample (NFHS reports are focused on women’s health factors), single-day measurement of BP which is not recommended to confirm the diagnosis of hypertension, use of older definition of hypertension (SBP ≥140 and/or DBP ≥90 mmHg) instead of more recently suggested criteria (SBP ≥130 and/or DBP ≥80 mmHg). In the present study we do not yet have data on hypertension awareness, awareness and control status and this is also a limitation. Data on pre-hypertension, which is more prevalent than hypertension in India, and should be the focus of prevention strategies, are not yet available. There is a need for more primary data on hypertension determinants to explain regional variations including social and cultural determinants of physical activity and diet. Data on district-level availability and access to healthcare and availability of drug-therapies to individuals, especially women, with hypertension are not available. Finally, all these data need replication for external validation using data from concurrent studies including the ICMR-INDIAB and other national surveys.

## Conclusions

It has been argued that phase transition in the human progress has been brought about by tectonic shifts in development with technological progress in every corner of the globe. This has lifted most societies out of Malthusian trap but aspects of local geography, culture and institutions are also important in this transition.^33^ Our study shows that the escape from poverty and illiteracy among women is associated with greater hypertension prevalence in India and forecasts a short-term scenario with more hypertension with better social development. We hope that a socioeconomic tipping point shall arrive soon where hypertension is more in lower socioeconomic status individuals.^34^ India and most low-middle income and low-income countries have significantly higher adverse cardiovascular events and mortality from hypertension.^35^ Reversal of this trend is possible using well-tried evidence-based interventions at the level of population and individuals cross the life course. The creation of an egalitarian society with a focus on various components of inequality,^34^ availability of non-communicable disease-focused healthcare,^36^ and population empowerment,^36^ would lead to lower hypertension among the more developed communities and regions of the country.

## Data Availability

Data from NFHS-5 are available at the National Family Health Survey website at http://rchiips.org/nfhs/factsheet_NFHS-5.shtml

## CONTRIBUTORS

RG and KG conceptualized the study. Data analyses and estimate generation were done by KG and SS. Maps were generated by KG. RG drafted the first iteration of the manuscript. RG, KG, SS and RKD made substantial contributions to critical review, editing and revision of the manuscript. All the authors approved the final version of the manuscript. RG, KG and SS accessed and verified the data underlying the study. All authors had full access to all the data in the study and had final responsibility for the decision to submit for publication.

## DATA SHARING

All the data used for the manuscript are freely available at the National Family Health Survey website at http://rchiips.org/nfhs/.

## ACKNOWLEDGEMENTS

No specific funding was available for this article.

