## Supplementary Tables for "District Level Variation in Hypertension Epidemiology in India and Influence of Social Determinants: National Family Health Survey-5"

Supplementary Tables (1-35): Prevalence of hypertension in various districts in each state of India

Supplementary Figures: 1-4

**Supplementary** **Table 1: UN Sustainable Development Goals (SDGs) important for cardiovascular health**

| **SDG Number** | **SDG domain** | **WHO response** | **Relevance to CVD prevention & control** |
| --- | --- | --- | --- |
| 1 | No poverty | Prioritizing the health needs of the poor | ++++ |
| 2 | Zero hunger | Addressing the causes and consequences of all forms of malnutrition | +++ |
| 3 | Good health and well being | Ensure healthy lives and promote wellbeing for all at all ages | ++++ |
| 4 | Quality education | Supporting high quality education for all to improve health and health equity | ++++ |
| 5 | Gender equality | Fighting gender inequality including violence against women | ++ |
| 6 | Clean water and sanitation | Preventing disease through safe water and sanitation for all | + |
| 7 | Affordable and clean energy | Promoting sustainable energy for healthy homes and lives | ++ |
| 8 | Decent work and economic growth | Promoting health employment as a driver of inclusive economic growth | ++ |
| 9 | Industry, innovation and infrastructure | Promoting national research and development capacity and manufacturing of affordable essential medical products | +++ |
| 10 | Reduced inequalities | Ensuring equitable access to health services through universal health coverage based on strong primary care | +++ |
| 11 | Sustainable cities and communities | Fostering healthier cities through urban planning and cleaner air and safer and more active living | +++ |
| 12 | Responsible consumption and production | Promoting responsible consumption of medicines to combat antibiotic resistance (or overmedication) | ++ |
| 13 | Climate action | Protective health from climate risks and promoting health through low-carbon development | +++ |
| 14 | Life below water | Supporting the restoration of fish stocks to improve safe and diversified healthy diets | +++ |
| 15 | Life on land | Promoting health and preventing disease through healthy natural environments | +++ |
| 16 | Peace, justice and strong institutions | Empowering strong local institutions to develop, implement, monitor and account for ambitious SDG responses | +++ |
| 17 | Partnerships for the goals | Mobilizing partners to monitor and attain health related SDGs | +++ |

**Supplementary** **Tables 2:** Prevalence of hypertension in various districts of Andhra Pradesh

|  | Sample size | | Age-standardized hypertension  prevalence (%) | |
| --- | --- | --- | --- | --- |
|  | Women | Men | Women | Men |
| Andhra Pradesh |  |  |  |  |
| Anantapur | 868 | 146 | 23.4 | 26.7 |
| Chittoor | 828 | 121 | 22.7 | 27.1 |
| East Godavari | 824 | 105 | 29.0 | 32.2 |
| Guntur | 807 | 93 | 25.8 | 31.3 |
| Krishna | 820 | 119 | 24.0 | 28.5 |
| Kurnool | 908 | 139 | 27.0 | 31.8 |
| Prakasam | 689 | 95 | 27.8 | 33.0 |
| Sri Potti Sriramulu Nello | 922 | 132 | 23.0 | 25.1 |
| Visakhapatnam | 818 | 112 | 23.9 | 29.2 |
| West Godavari | 841 | 122 | 28.8 | 31.4 |
| Y.S.R. | 1017 | 140 | 23.2 | 27.3 |
| Srikakulam | 780 | 100 | 22.1 | 22.9 |
| Vizianagaram | 853 | 134 | 25.2 | 25.1 |
| Mean |  |  | 25.1 | 28.6 |
| Median |  |  | 24.0 | 28.5 |
| Maximum |  |  | 29.0 | 33.0 |
| Minimum |  |  | 22.1 | 22.9 |
| Interquartile Interval |  |  | 3.76 | 4.75 |

Supplementary Table 3: Prevalence of hypertension in various districts of Arunachal Pradesh

| States | Sample size | | Age-standardized hypertension  prevalence (%) | |
| --- | --- | --- | --- | --- |
|  | Women | Men | Women | Men |
| Arunachal Pradesh |  |  |  |  |
| Anjaw | 828 | 125 | 31.1 | 42.6 |
| Changlang | 1168 | 189 | 22.8 | 27.5 |
| Dibang Valley | 868 | 134 | 32.2 | 45.4 |
| East Kameng | 1002 | 130 | 19.5 | 29.0 |
| East Siang | 1157 | 161 | 29.7 | 38.1 |
| Kra Daadi | 524 | 75 | 19.4 | 26.1 |
| Kurung Kumey | 650 | 63 | 19.2 | 26.0 |
| Lohit | 1129 | 193 | 22.1 | 26.6 |
| Longding | 935 | 149 | 20.5 | 31.1 |
| Lower Dibang Valley | 1090 | 173 | 25.6 | 38.1 |
| Lower Subansiri | 1094 | 167 | 25.7 | 36.6 |
| Namsai | 1156 | 171 | 27.1 | 28.1 |
| Papum Pare | 1017 | 142 | 20.6 | 32.7 |
| Siang | 1106 | 154 | 24.4 | 31.5 |
| Tawang | 924 | 137 | 30.3 | 36.4 |
| Tirap | 934 | 138 | 25.2 | 31.6 |
| Upper Siang | 901 | 133 | 26.1 | 31.7 |
| Upper Subansiri | 1092 | 147 | 25.4 | 37.5 |
| West Kameng | 1122 | 167 | 19.8 | 30.7 |
| West Siang | 1068 | 133 | 32.7 | 41.1 |
| Mean |  |  | 24.2 | 32.6 |
| Median |  |  | 25.3 | 31.6 |
| Maximum |  |  | 32.2 | 45.4 |
| Minimum |  |  | 7.2 | 9.1 |
| Interquartile Interval |  |  | 7.17 | 9.06 |

Supplementary Table 4 : Prevalence of hypertension in various districts of Assam

| States | Sample size | | Age-standardized hypertension  prevalence (%) | |
| --- | --- | --- | --- | --- |
|  | Women | Men | Women | Men |
| **Assam** |  |  |  |  |
| Baksa | 1175 | 156 | 24.5 | 25.0 |
| Barpeta | 1163 | 159 | 17.9 | 16.7 |
| Biswanath | 1000 | 135 | 23.5 | 28.4 |
| Bongaigaon | 1092 | 169 | 13.6 | 15.9 |
| Cachar | 1110 | 152 | 17.0 | 15.4 |
| Charaideo | 1117 | 166 | 18.2 | 17.9 |
| Chirang | 1076 | 148 | 20.8 | 25.6 |
| Darrang | 1053 | 176 | 16.7 | 14.5 |
| Dhemaji | 989 | 148 | 16.0 | 24.1 |
| Dhubri | 1017 | 128 | 18.2 | 19.0 |
| Dibrugarh | 1086 | 161 | 18.1 | 21.4 |
| Dima Hasao | 1001 | 149 | 17.1 | 23.5 |
| Goalpara | 1158 | 164 | 16.8 | 17.9 |
| Golaghat | 1028 | 129 | 20.5 | 22.0 |
| Hailakandi | 1057 | 130 | 21.8 | 24.4 |
| Hojai | 1027 | 148 | 21.0 | 25.2 |
| Jorhat | 1019 | 148 | 25.9 | 26.3 |
| Kamrup | 1021 | 138 | 22.2 | 19.2 |
| Kamrup Metropolitan | 926 | 165 | 17.0 | 20.3 |
| Karbi Anglong | 1042 | 156 | 20.1 | 22.8 |
| Karimganj | 1170 | 160 | 16.5 | 17.2 |
| Kokrajhar | 1052 | 144 | 17.8 | 19.4 |
| Lakhimpur | 957 | 140 | 20.8 | 25.0 |
| Majuli | 1052 | 146 | 19.7 | 23.3 |
| Morigaon | 1071 | 136 | 20.1 | 20.5 |
| Nagaon | 1122 | 169 | 16.4 | 17.0 |
| Nalbari | 1011 | 145 | 22.5 | 24.9 |
| Sivasagar | 1020 | 146 | 22.0 | 26.0 |
| Sonitpur | 1067 | 148 | 24.8 | 24.5 |
| South Salmara Mancachar | 1085 | 162 | 15.5 | 16.5 |
| Tinsukia | 1062 | 155 | 17.1 | 19.3 |
| Udalguri | 1088 | 151 | 19.7 | 22.6 |
| West Karbi Anglong | 1065 | 146 | 17.0 | 22.2 |
| Mean |  |  | 19.3 | 21.3 |
| Median |  |  | 18.2 | 22.0 |
| Maximum |  |  | 25.9 | 28.4 |
| Minimum |  |  | 13.6 | 14.5 |
| Interquartile Interval |  |  | 3.96 | 6.59 |

Supplementary Table 5 : Prevalence of hypertension in various districts of Bihar

| States | Sample size | | Age-standardized hypertension  prevalence (%) | |
| --- | --- | --- | --- | --- |
|  | Women | Men | Women | Men |
| **Bihar** |  |  |  |  |
| Araria | 1122 | 124 | 12.1 | 15.8 |
| Arwal | 1185 | 133 | 12.9 | 14.0 |
| Aurangabad | 1269 | 161 | 16.2 | 15.3 |
| Banka | 1098 | 147 | 17.4 | 18.8 |
| Begusarai | 1170 | 165 | 13.9 | 17.0 |
| Bhagalpur | 1154 | 179 | 13.3 | 14.7 |
| Bhojpur | 1213 | 181 | 13.9 | 14.5 |
| Buxar | 1295 | 158 | 13.9 | 10.5 |
| Darbhanga | 1053 | 100 | 17.5 | 23.5 |
| Gaya | 1153 | 147 | 16.6 | 18.8 |
| Gopalganj | 1139 | 96 | 20.3 | 22.8 |
| Jamui | 1172 | 119 | 12.6 | 14.8 |
| Jehanabad | 1072 | 156 | 13.4 | 12.0 |
| Kaimur (Bhabua) | 1167 | 146 | 12.3 | 15.0 |
| Katihar | 945 | 107 | 17.0 | 17.8 |
| Khagaria | 1072 | 125 | 13.0 | 16.1 |
| Kishanganj | 1116 | 111 | 17.4 | 22.7 |
| Lakhisarai | 1222 | 147 | 12.3 | 13.8 |
| Madhepura | 1055 | 127 | 10.4 | 14.5 |
| Madhubani | 976 | 83 | 18.2 | 23.4 |
| Munger | 1100 | 157 | 16.1 | 18.3 |
| Muzaffarpur | 1095 | 94 | 18.1 | 24.0 |
| Nalanda | 1151 | 146 | 13.4 | 14.0 |
| Nawada | 1239 | 121 | 14.5 | 18.5 |
| Pashchim Champaran | 1070 | 109 | 18.1 | 22.6 |
| Patna | 942 | 143 | 16.1 | 14.8 |
| Purba Champaran | 1142 | 87 | 16.2 | 23.6 |
| Purnia | 973 | 133 | 18.7 | 22.5 |
| Rohtas | 1302 | 148 | 15.2 | 16.0 |
| Saharsa | 960 | 110 | 14.9 | 15.5 |
| Samastipur | 1061 | 118 | 15.8 | 16.4 |
| Saran | 1229 | 137 | 16.7 | 19.3 |
| Sheikhpura | 1160 | 132 | 16.3 | 16.1 |
| Sheohar | 886 | 104 | 15.5 | 20.8 |
| Sitamarhi | 1004 | 116 | 17.8 | 17.9 |
| Siwan | 1256 | 95 | 19.6 | 29.6 |
| Supaul | 1153 | 112 | 11.6 | 14.1 |
| Vaishali | 1112 | 123 | 17.4 | 19.6 |
| Mean |  |  | 15.4 | 17.9 |
| Median |  |  | 16.0 | 16.7 |
| Maximum |  |  | 20.3 | 29.6 |
| Minimum |  |  | 10.4 | 10.5 |
| Interquartile Interval |  |  | 4.04 | 5.72 |

Supplementary Table 6 : Prevalence of hypertension in various districts of Chandigarh

| States | Sample size | | Age-standardized hypertension  prevalence (%) | |
| --- | --- | --- | --- | --- |
|  | Women | Men | Women | Men |
| Chandigarh | 746 | 104 | 25.0 | 30.6 |

Supplementary Table 7 : Prevalence of hypertension in various districts of Chhatisgarh

| States | Sample size | | Age-standardized hypertension prevalence (%) | |
| --- | --- | --- | --- | --- |
|  | Women | Men | Women | Men |
| **Chhatisgarh** |  |  |  |  |
| Balod | 1006 | 164 | 25.1 | 27.7 |
| Baloda Bazar | 1116 | 169 | 22.6 | 27.9 |
| Balrampur | 995 | 128 | 25.2 | 31.2 |
| Bastar | 1036 | 143 | 23.7 | 24.1 |
| Bemetara | 1060 | 134 | 26.9 | 27.6 |
| Bijapur | 1087 | 166 | 15.0 | 21.6 |
| Bilaspur | 973 | 173 | 24.4 | 28.6 |
| Dantewada | 1121 | 162 | 19.1 | 23.7 |
| Dhamtari | 1152 | 151 | 21.0 | 24.9 |
| Durg | 1112 | 184 | 22.6 | 26.5 |
| Gariyaband | 1044 | 159 | 22.6 | 25.6 |
| Janjgir Champa | 1046 | 149 | 21.6 | 28.3 |
| Jashpur | 945 | 136 | 28.5 | 34.4 |
| Kabeerdham | 1037 | 151 | 24.0 | 26.5 |
| Kodagaon | 1165 | 148 | 21.2 | 21.6 |
| Korba | 1029 | 173 | 23.9 | 32.7 |
| Koriya | 967 | 133 | 25.7 | 30.9 |
| Mahasamund | 1073 | 163 | 17.9 | 21.4 |
| Mungeli | 963 | 119 | 26.2 | 29.2 |
| Narayanpur | 1146 | 164 | 18.8 | 21.1 |
| Raigarh | 990 | 154 | 26.1 | 31.9 |
| Raipur | 1173 | 182 | 17.7 | 20.6 |
| Rajnandgaon | 1030 | 172 | 27.7 | 28.3 |
| Sukma | 1218 | 173 | 14.5 | 18.1 |
| Surajpur | 1024 | 135 | 25.0 | 30.9 |
| Surguja | 950 | 133 | 28.5 | 37.5 |
| Uttar Bastar Kanker | 1010 | 156 | 27.0 | 30.2 |
| Mean |  |  | 23.0 | 27.1 |
| Median |  |  | 23.9 | 27.7 |
| Maximum |  |  | 28.5 | 37.5 |
| Minimum |  |  | 14.5 | 18.1 |
| Interquartile Interval |  |  | 4.84 | 6.65 |

Supplementary Table 8: Prevalence of hypertension in various districts of Goa

| States | Sample size | | Age-standardized hypertension | |
| --- | --- | --- | --- | --- |
|  |  |  | prevalence (%) | |
|  | Women | Men | Women | Men |
| **Goa** |  |  |  |  |
| North Goa | 975 | 148 | 27.3 | 28.3 |
| South Goa | 1055 | 165 | 27.7 | 25.0 |
| Mean |  |  | 27.5 | 26.7 |
| Median |  |  | 27.5 | 26.7 |
| Maximum |  |  | 27.7 | 28.3 |
| Minimum |  |  | 27.3 | 25.0 |
| Interquartile Interval |  |  | 0.25 | 1.62 |

Supplementary Table 9 : Prevalence of hypertension in various districts of Gujarat

| States | Sample size | | Age-standardized hypertension  prevalence (%) | |
| --- | --- | --- | --- | --- |
|  | Women | Men | Women | Men |
| **Gujarat** |  |  |  |  |
| Ahmadabad | 1010 | 155 | 18.5 | 16.2 |
| Amreli | 936 | 127 | 18.0 | 11.5 |
| Anand | 914 | 163 | 26.6 | 26.9 |
| Aravali | 1078 | 187 | 21.0 | 22.0 |
| Banas Kantha | 1049 | 157 | 14.3 | 15.7 |
| Bharuch | 919 | 124 | 23.7 | 24.9 |
| Bhavnagar | 950 | 145 | 20.1 | 18.0 |
| Botad | 1032 | 149 | 12.5 | 12.0 |
| Chhota Udaipur | 1035 | 173 | 19.4 | 17.5 |
| Devbhumi Dwarka | 1170 | 198 | 18.1 | 17.3 |
| Dohad | 1221 | 158 | 18.2 | 17.7 |
| Gandhinagar | 919 | 151 | 23.9 | 29.3 |
| Gir Somnath | 1158 | 185 | 14.0 | 12.3 |
| Jamnagar | 1042 | 165 | 18.1 | 15.0 |
| Junagadh | 963 | 164 | 23.3 | 20.8 |
| Kachchh | 1026 | 181 | 19.1 | 18.2 |
| Kheda | 1029 | 183 | 24.0 | 24.5 |
| Mahesana | 883 | 136 | 22.6 | 27.0 |
| Mahisagar | 1017 | 168 | 16.3 | 14.7 |
| Morbi | 1119 | 182 | 18.4 | 15.1 |
| Narmada | 1021 | 155 | 19.2 | 20.8 |
| Navsari | 991 | 162 | 23.7 | 22.1 |
| Panch Mahals | 1101 | 196 | 22.5 | 27.6 |
| Patan | 1011 | 169 | 16.3 | 16.3 |
| Porbandar | 1018 | 169 | 15.8 | 16.2 |
| Rajkot | 979 | 159 | 17.8 | 14.5 |
| Sabar Kantha | 1018 | 152 | 24.0 | 26.1 |
| Surat | 986 | 195 | 23.4 | 22.6 |
| Surendranagar | 931 | 143 | 17.1 | 14.7 |
| Tapi | 991 | 144 | 25.2 | 28.0 |
| The Dangs | 901 | 130 | 22.9 | 24.1 |
| Vadodara | 894 | 167 | 24.3 | 23.8 |
| Valsad | 1031 | 159 | 23.8 | 29.3 |
| Mean |  |  | 20.2 | 20.1 |
| Median |  |  | 19.4 | 18.2 |
| Maximum |  |  | 26.6 | 29.3 |
| Minimum |  |  | 12.5 | 11.5 |
| Interquartile Interval |  |  | 5.71 | 8.83 |

Supplementary Table 10: Prevalence of hypertension in various districts of Haryana

| States | Sample size | | Age-standardized hypertension | |
| --- | --- | --- | --- | --- |
|  |  |  | prevalence (%) | |
|  | Women | Men | Women | Men |
| **Haryana** |  |  |  |  |
| Ambala | 1139 | 200 | 28.8 | 31.2 |
| Bhiwani | 1046 | 143 | 18.7 | 24.8 |
| Charkhi Dadri | 960 | 113 | 20.6 | 22.9 |
| Faridabad | 1033 | 184 | 23.3 | 26.2 |
| Fatehabad | 899 | 145 | 17.5 | 20.1 |
| Gurgaon | 849 | 98 | 16.5 | 22.4 |
| Hisar | 1016 | 152 | 19.4 | 28.6 |
| Jhajjar | 911 | 142 | 21.0 | 26.8 |
| Jind | 1053 | 159 | 16.5 | 21.9 |
| Kaithal | 872 | 97 | 16.9 | 20.4 |
| Karnal | 1237 | 202 | 17.6 | 21.2 |
| Kurukshetra | 1140 | 184 | 30.4 | 34.4 |
| Mahendragarh | 997 | 164 | 21.6 | 25.8 |
| Mewat | 1078 | 159 | 15.5 | 19.2 |
| Palwal | 1166 | 191 | 21.9 | 23.2 |
| Panchkula | 751 | 83 | 24.3 | 26.2 |
| Panipat | 1004 | 143 | 21.6 | 25.6 |
| Rewari | 1021 | 150 | 24.1 | 26.5 |
| Rohtak | 978 | 139 | 20.7 | 24.5 |
| Sirsa | 588 | 28 | 19.9 | 26.8 |
| Sonipat | 1017 | 156 | 20.0 | 21.9 |
| Yamunanagar | 1154 | 192 | 28.7 | 28.8 |
| Mean |  |  | 21.2 | 25.0 |
| Median |  |  | 20.7 | 25.2 |
| Maximum |  |  | 30.4 | 34.4 |
| Minimum |  |  | 15.5 | 19.2 |
| Interquartile Interval |  |  | 5.08 | 4.67 |

Supplementary Table 11 : Prevalence of hypertension in various districts of Himachal Pradesh

| States | Sample size | | Age-standardized hypertension | |
| --- | --- | --- | --- | --- |
|  |  |  | prevalence (%) | |
|  | Women | Men | Women | Men |
| **Himachal Pradesh** |  |  |  |  |
| Bilaspur | 896 | 117 | 22.1 | 20.3 |
| Chamba | 931 | 109 | 17.8 | 23.2 |
| Hamirpur | 797 | 87 | 28.1 | 35.2 |
| Kangra | 930 | 123 | 24.9 | 26.6 |
| Kinnaur | 707 | 143 | 18.5 | 22.8 |
| Kullu | 892 | 128 | 17.4 | 20.6 |
| Lahul & Spiti | 698 | 113 | 15.0 | 19.2 |
| Mandi | 878 | 112 | 20.8 | 23.8 |
| Shimla | 864 | 126 | 19.9 | 23.5 |
| Sirmaur | 1044 | 167 | 19.1 | 20.8 |
| Solan | 855 | 148 | 21.7 | 22.3 |
| Una | 876 | 104 | 27.1 | 27.2 |
| Mean |  |  | 21.0 | 23.8 |
| Median |  |  | 20.4 | 23.0 |
| Maximum |  |  | 28.1 | 35.2 |
| Minimum |  |  | 15.0 | 19.2 |
| Interquartile Interval |  |  | 4.47 | 3.79 |

Supplementary Table 12 : Prevalence of hypertension in various districts of Jammu & Kashmir

| States | Sample size | | Age-standardized hypertension prevalence (%) | |
| --- | --- | --- | --- | --- |
|  | Women | Men | Women | Men |
| Jammu & Kashmir |  |  |  |  |
| Anantnag | 1102 | 125 | 20.0 | 19.4 |
| Baramula | 1329 | 166 | 23.7 | 19.0 |
| Kulgam | 1190 | 139 | 25.8 | 19.9 |
| Shupiyan | 1135 | 148 | 15.1 | 14.2 |
| Reasi | 1100 | 140 | 15.4 | 17.6 |
| Rajouri | 1183 | 181 | 16.5 | 13.8 |
| Jammu | 1050 | 125 | 20.5 | 22.6 |
| Srinagar | 1030 | 138 | 20.1 | 18.1 |
| Punch | 1182 | 168 | 17.9 | 16.3 |
| Kupwara | 1233 | 175 | 18.1 | 18.9 |
| Samba | 1121 | 150 | 21.1 | 21.5 |
| Bandipore | 1263 | 177 | 21.4 | 17.4 |
| Kathua | 1064 | 149 | 18.9 | 23.6 |
| Kishtwar | 1158 | 152 | 13.2 | 13.3 |
| Doda | 1000 | 141 | 23.1 | 21.5 |
| Pulwama | 1155 | 151 | 22.6 | 19.1 |
| Ramban | 1192 | 182 | 17.5 | 15.7 |
| Ganderbal | 1202 | 158 | 21.5 | 19.1 |
| Udhampur | 1148 | 150 | 20.1 | 18.3 |
| Badgam | 1200 | 172 | 19.9 | 18.6 |
| Mean |  |  | 19.6 | 18.4 |
| Median |  |  | 20.0 | 18.7 |
| Maximum |  |  | 25.8 | 23.6 |
| Minimum |  |  | 13.2 | 13.3 |
| Interquartile Interval |  |  | 3.66 | 2.41 |

Supplementary Table 13: Prevalence of hypertension in various districts of Jharkhand

| States | Sample size | | Age-standardized hypertension  prevalence (%) | |
| --- | --- | --- | --- | --- |
|  | Women | Men | Women | Men |
| **Jharkhand** |  |  |  |  |
| Bokaro | 1127 | 157 | 21.5 | 27.5 |
| Chatra | 1154 | 120 | 16.0 | 22.1 |
| Deoghar | 1066 | 136 | 14.2 | 18.5 |
| Dhanbad | 1181 | 166 | 18.5 | 24.9 |
| Dumka | 958 | 122 | 14.1 | 20.0 |
| Garhwa | 1060 | 128 | 18.3 | 23.4 |
| Giridih | 1124 | 114 | 17.4 | 23.9 |
| Godda | 913 | 109 | 18.6 | 20.2 |
| Gumla | 1113 | 176 | 15.1 | 19.9 |
| Hazaribagh | 1189 | 95 | 18.8 | 30.8 |
| Jamtara | 1073 | 144 | 17.8 | 21.9 |
| Khunti | 1103 | 142 | 11.7 | 14.2 |
| Kodarma | 1227 | 142 | 13.8 | 20.6 |
| Latehar | 1148 | 161 | 15.3 | 22.2 |
| Lohardaga | 1107 | 145 | 17.5 | 20.2 |
| Pakur | 966 | 104 | 17.1 | 20.2 |
| Palamu | 1204 | 143 | 18.3 | 24.3 |
| Pashchimi Singhbhum | 1076 | 145 | 17.8 | 19.5 |
| Purbi Singhbhum | 1133 | 177 | 19.8 | 22.4 |
| Ramgarh | 1123 | 178 | 21.1 | 27.4 |
| Ranchi | 1182 | 156 | 19.6 | 23.1 |
| Sahibganj | 1007 | 113 | 15.0 | 16.6 |
| Saraikela Kharsawan | 1126 | 183 | 15.6 | 17.2 |
| Simdega | 1135 | 158 | 20.9 | 26.7 |
| Mean |  |  | 17.2 | 22.0 |
| Median |  |  | 17.6 | 22.0 |
| Maximum |  |  | 21.5 | 30.8 |
| Minimum |  |  | 11.7 | 14.2 |
| Interquartile Interval |  |  | 3.36 | 3.94 |

Supplementary Table 14: Prevalence of hypertension in various districts of Karnataka

| States | Sample size | | Age-standardized hypertension | |
| --- | --- | --- | --- | --- |
|  |  |  | prevalence (%) | |
|  | Women | Men | Women | Men |
| **Karnataka** |  |  |  |  |
| Bagalkot | 1138 | 182 | 19.1 | 21.0 |
| Bangalore Rural | 956 | 142 | 26.7 | 30.8 |
| Bellary | 1103 | 156 | 21.2 | 20.9 |
| Bidar | 1181 | 172 | 21.2 | 24.1 |
| Bijapur | 1091 | 149 | 20.2 | 21.0 |
| Chamarajanagar | 957 | 136 | 28.8 | 29.9 |
| Chikkaballapura | 881 | 144 | 24.1 | 32.5 |
| Chikmagalur | 945 | 143 | 29.2 | 28.7 |
| Chitradurga | 953 | 134 | 29.7 | 30.6 |
| Dakshina Kannada | 987 | 149 | 30.1 | 29.9 |
| Davanagere | 973 | 133 | 24.8 | 29.5 |
| Dharwad | 1051 | 156 | 26.6 | 23.7 |
| Gadag | 1136 | 193 | 22.1 | 23.9 |
| Gulbarga | 1147 | 167 | 17.0 | 24.8 |
| Hassan | 979 | 136 | 28.1 | 29.5 |
| Haveri | 1060 | 141 | 21.4 | 24.4 |
| Kodagu | 885 | 139 | 31.5 | 33.3 |
| Kolar | 990 | 164 | 24.3 | 27.9 |
| Mandya | 844 | 127 | 30.1 | 29.8 |
| Mysore | 951 | 134 | 25.5 | 27.4 |
| Raichur | 1177 | 166 | 18.5 | 16.7 |
| Shimoga | 1033 | 161 | 27.2 | 28.7 |
| Udupi | 1065 | 145 | 31.2 | 32.5 |
| Uttara Kannada | 1000 | 164 | 26.9 | 25.1 |
| Yadgir | 1242 | 171 | 17.5 | 18.0 |
| Mean |  |  | 24.9 | 26.6 |
| Median |  |  | 25.5 | 27.9 |
| Maximum |  |  | 31.5 | 33.3 |
| Minimum |  |  | 17.0 | 16.7 |
| Interquartile Interval |  |  | 7.53 | 5.97 |

Supplementary Table 15 : Prevalence of hypertension in various districts of Kerela

| States | Sample size | | Age-standardized hypertension | |
| --- | --- | --- | --- | --- |
|  |  |  | prevalence (%) | |
|  | Women | Men | Women | Men |
| **Kerela** |  |  |  |  |
| Alappuzha | 685 | 111 | 32.1 | 36.2 |
| Ernakulam | 747 | 115 | 29.6 | 30.2 |
| Idukki | 710 | 121 | 34.8 | 31.8 |
| Kannur | 950 | 101 | 31.0 | 34.8 |
| Kasaragod | 945 | 116 | 26.1 | 26.0 |
| Kollam | 753 | 78 | 32.4 | 35.3 |
| Kottayam | 659 | 100 | 34.2 | 38.8 |
| Kozhikode | 825 | 107 | 29.6 | 28.4 |
| Malappuram | 1004 | 141 | 25.4 | 27.4 |
| Palakkad | 881 | 113 | 32.4 | 35.3 |
| Pathanamthitta | 625 | 92 | 42.1 | 41.9 |
| Thiruvananthapuram | 657 | 84 | 31.4 | 32.1 |
| Thrissur | 715 | 66 | 31.4 | 36.8 |
| Wayanad | 813 | 128 | 28.8 | 28.3 |
| Mean |  |  | 31.5 | 33.1 |
| Median |  |  | 31.4 | 33.5 |
| Maximum |  |  | 42.1 | 41.9 |
| Minimum |  |  | 25.4 | 26.0 |
| Interquartile Interval |  |  | 2.80 | 7.10 |

Supplementary Table 16 : Prevalence of hypertension in various districts of Madhya Pradesh

| States | Sample size | | Age-standardized hypertension | |
| --- | --- | --- | --- | --- |
|  |  |  | prevalence (%) | |
|  | Women | Men | Women | Men |
| Madhya Pradesh |  |  |  |  |
| Agar Malwa | 588 | 82 | 21.2 | 25.2 |
| Alirajpur | 1080 | 157 | 22.0 | 27.7 |
| Anuppur | 1020 | 187 | 24.3 | 26.4 |
| Ashoknagar | 862 | 96 | 16.3 | 19.8 |
| Balaghat | 860 | 137 | 22.3 | 21.9 |
| Barwani | 1236 | 189 | 21.1 | 25.0 |
| Betul | 1021 | 141 | 17.8 | 21.6 |
| Bhind | 956 | 133 | 15.2 | 17.0 |
| Bhopal | 337 | 38 | 22.4 | 24.9 |
| Burhanpur | 1139 | 197 | 13.9 | 15.9 |
| Chhatarpur | 1001 | 161 | 17.2 | 17.5 |
| Chhindwara | 1009 | 128 | 21.7 | 22.6 |
| Damoh | 988 | 160 | 22.6 | 21.4 |
| Datia | 1015 | 145 | 15.3 | 19.7 |
| Dewas | 1107 | 188 | 24.4 | 28.6 |
| Dhar | 679 | 101 | 19.9 | 19.8 |
| Dindori | 1047 | 163 | 25.2 | 27.7 |
| Guna | 1165 | 162 | 17.0 | 18.2 |
| Gwalior | 958 | 123 | 19.9 | 25.1 |
| Harda | 1207 | 204 | 21.7 | 20.9 |
| Hoshangabad | 1042 | 162 | 24.0 | 29.9 |
| Indore | 1071 | 188 | 20.7 | 21.3 |
| Jabalpur | 216 | 36 | 22.7 | 20.1 |
| Jhabua | 1037 | 140 | 23.5 | 23.7 |
| Katni | 777 | 101 | 18.5 | 20.8 |
| Khandwa (East Nimar) | 562 | 17 | 21.3 | 17.6 |
| Khargone (West Nimar) | 1173 | 180 | 21.2 | 24.9 |
| Mandla | 1049 | 153 | 24.9 | 30.4 |
| Mandsaur | 1015 | 173 | 29.6 | 36.2 |
| Morena | 1079 | 137 | 14.9 | 19.1 |
| Narsimhapur | 1110 | 160 | 23.2 | 24.4 |
| Neemuch | 1178 | 181 | 26.0 | 28.5 |
| Panna | 992 | 152 | 16.1 | 14.4 |
| Raisen | 474 | 52 | 19.0 | 21.2 |
| Rajgarh | 1020 | 144 | 24.4 | 24.0 |
| Ratlam | 1087 | 158 | 21.3 | 24.0 |
| Rewa | 927 | 83 | 17.6 | 20.1 |
| Sagar | 1014 | 174 | 25.3 | 26.4 |
| Satna | 689 | 79 | 16.2 | 16.3 |
| Sehore | 1088 | 168 | 26.0 | 26.6 |
| Seoni | 989 | 136 | 21.1 | 25.7 |
| Shahdol | 860 | 127 | 19.8 | 23.2 |
| Shajapur | 1226 | 184 | 23.3 | 26.1 |
| Sheopur | 997 | 119 | 19.2 | 25.7 |
| Shivpuri | 790 | 115 | 13.7 | 15.7 |
| Sidhi | 1148 | 136 | 17.5 | 26.0 |
| Singrauli | 643 | 64 | 18.9 | 25.9 |
| Tikamgarh | 620 | 82 | 14.4 | 12.9 |
| Ujjain | 1108 | 194 | 23.3 | 25.0 |
| Umaria | 1135 | 172 | 20.9 | 22.1 |
| Vidisha | 1019 | 166 | 17.2 | 18.1 |
| Mean |  |  | 20.5 | 22.8 |
| Median |  |  | 21.1 | 23.2 |
| Maximum |  |  | 29.6 | 36.2 |
| Minimum |  |  | 13.7 | 12.9 |
| Interquartile Interval |  |  | 5.70 | 5.97 |

Supplementary Table 17 : Prevalence of hypertension in various districts of Maharashtra

| States | Sample size | | Age-standardized hypertension | |
| --- | --- | --- | --- | --- |
|  |  |  | prevalence (%) | |
|  | Women | Men | Women | Men |
| **Maharashtra** |  |  |  |  |
| Ahmadnagar | 993 | 139 | 23.6 | 25.3 |
| Akola | 1098 | 202 | 19.4 | 19.1 |
| Amravati | 1060 | 180 | 19.8 | 18.7 |
| Aurangabad | 1011 | 147 | 20.8 | 20.9 |
| Bhandara | 920 | 152 | 21.8 | 19.3 |
| Bid | 864 | 146 | 26.4 | 24.1 |
| Buldana | 1055 | 189 | 20.1 | 20.5 |
| Chandrapur | 965 | 175 | 20.7 | 20.6 |
| Dhule | 883 | 137 | 21.4 | 25.1 |
| Gadchiroli | 915 | 164 | 17.0 | 17.2 |
| Gondiya | 938 | 167 | 21.8 | 18.9 |
| Hingoli | 1121 | 173 | 17.4 | 17.4 |
| Jalgaon | 881 | 155 | 20.7 | 24.4 |
| Jalna | 937 | 145 | 21.8 | 24.8 |
| Kolhapur | 986 | 150 | 31.0 | 33.2 |
| Latur | 1006 | 160 | 18.6 | 19.6 |
| Mumbai | 779 | 127 | 21.0 | 23.3 |
| Mumbai Suburban | 552 | 64 | 26.6 | 28.0 |
| Nagpur | 1063 | 169 | 21.3 | 18.3 |
| Nanded | 1026 | 164 | 16.7 | 17.3 |
| Nandurbar | 1040 | 171 | 23.5 | 24.6 |
| Nashik | 1023 | 155 | 22.5 | 26.1 |
| Osmanabad | 916 | 147 | 23.5 | 26.2 |
| Palghar | 941 | 131 | 23.4 | 22.4 |
| Parbhani | 888 | 158 | 21.0 | 21.6 |
| Pune | 832 | 140 | 22.4 | 25.8 |
| Raigarh | 918 | 172 | 24.8 | 26.4 |
| Ratnagiri | 807 | 127 | 32.4 | 33.9 |
| Sangli | 966 | 152 | 30.1 | 32.0 |
| Satara | 978 | 149 | 27.2 | 29.8 |
| Sindhudurg | 722 | 115 | 36.6 | 37.9 |
| Solapur | 1003 | 153 | 25.7 | 26.2 |
| Thane | 756 | 91 | 24.5 | 29.7 |
| Wardha | 925 | 158 | 17.1 | 14.0 |
| Washim | 986 | 196 | 21.9 | 21.0 |
| Yavatmal | 1001 | 177 | 17.2 | 18.2 |
| Mean |  |  | 22.8 | 23.6 |
| Median |  |  | 21.8 | 23.7 |
| Maximum |  |  | 36.6 | 37.9 |
| Minimum |  |  | 16.7 | 14.0 |
| Interquartile Interval |  |  | 4.07 | 6.94 |

Supplementary Table 18: Prevalence of hypertension in various districts of Manipur

| States | Sample size | | Age-standardized hypertension | |
| --- | --- | --- | --- | --- |
|  |  |  | prevalence (%) | |
|  | Women | Men | Women | Men |
| **Manipur** |  |  |  |  |
| Bishnupur | 1043 | 140 | 23.6 | 31.3 |
| Chandel | 757 | 105 | 21.7 | 28.2 |
| Churachandpur | 872 | 119 | 16.3 | 25.2 |
| Imphal East | 1021 | 161 | 21.9 | 36.0 |
| Imphal West | 893 | 129 | 27.5 | 38.4 |
| Senapati | 837 | 120 | 18.7 | 28.2 |
| Tamenglong | 839 | 133 | 21.5 | 29.4 |
| Thoubal | 1069 | 149 | 23.2 | 33.4 |
| Ukhrul | 711 | 106 | 22.3 | 25.0 |
| Mean |  |  | 21.8 | 30.6 |
| Median |  |  | 21.9 | 29.4 |
| Maximum |  |  | 27.5 | 38.4 |
| Minimum |  |  | 16.3 | 25.0 |
| Interquartile Interval |  |  | 1.73 | 5.25 |

Supplementary Table 19 : Prevalence of hypertension in various districts of Meghalaya

| States | Sample size | | Age-standardized hypertension | |
| --- | --- | --- | --- | --- |
|  |  |  | prevalence (%) | |
|  | Women | Men | Women | Men |
| **Meghalaya** |  |  |  |  |
| East Garo Hills | 1280 | 198 | 15.3 | 23.2 |
| East Jantia Hills | 1311 | 200 | 18.7 | 17.2 |
| East Khasi Hills | 1066 | 131 | 24.5 | 25.0 |
| North Garo Hills | 1283 | 180 | 20.0 | 25.4 |
| Ribhoi | 1238 | 145 | 13.5 | 16.0 |
| South Garo Hills | 1088 | 143 | 15.8 | 22.8 |
| South West Garo Hills | 1161 | 140 | 13.6 | 20.0 |
| South West Khasi Hills | 1203 | 196 | 18.5 | 19.0 |
| West Garo Hills | 1090 | 158 | 17.4 | 24.7 |
| West Jaintia Hills | 1172 | 155 | 18.0 | 15.5 |
| West Khasi Hills | 1197 | 178 | 16.0 | 16.4 |
| Mean |  |  | 17.4 | 20.5 |
| Median |  |  | 17.4 | 20.0 |
| Maximum |  |  | 24.5 | 25.4 |
| Minimum |  |  | 13.5 | 15.5 |
| Interquartile Interval |  |  | 3.04 | 7.11 |

Supplementary Table 20 : Prevalence of hypertension in various districts of Mizoram

| States | Sample size | | Age-standardized hypertension | |
| --- | --- | --- | --- | --- |
|  |  |  | prevalence (%) | |
|  | Women | Men | Women | Men |
| **Mizoram** |  |  |  |  |
| Aizawl | 894 | 115 | 23.4 | 33.6 |
| Champhai | 806 | 150 | 16.1 | 19.6 |
| Kolasib | 926 | 145 | 16.5 | 24.3 |
| Lawngtlai | 975 | 153 | 13.0 | 20.3 |
| Lunglei | 973 | 128 | 13.9 | 20.0 |
| Mamit | 898 | 146 | 14.3 | 21.0 |
| Saiha | 890 | 134 | 13.0 | 22.9 |
| Serchhip | 917 | 134 | 11.6 | 17.7 |
| Mean |  |  | 15.2 | 22.4 |
| Median |  |  | 14.1 | 20.7 |
| Maximum |  |  | 23.4 | 33.6 |
| Minimum |  |  | 11.6 | 17.7 |
| Interquartile Interval |  |  | 3.17 | 3.30 |

Supplementary Table 21 : Prevalence of hypertension in various districts of Nagaland

| States | Sample size | | Age-standardized hypertension | |
| --- | --- | --- | --- | --- |
|  |  |  | prevalence (%) | |
|  | Women | Men | Women | Men |
| **Nagaland** |  |  |  |  |
| Dimapur | 1053 | 155 | 18.6 | 20.8 |
| Kiphire | 799 | 125 | 15.3 | 22.0 |
| Kohima | 817 | 132 | 25.7 | 31.1 |
| Longleng | 873 | 132 | 22.6 | 30.0 |
| Mokokchung | 892 | 118 | 23.2 | 28.9 |
| Mon | 832 | 123 | 20.4 | 35.7 |
| Peren | 943 | 148 | 19.8 | 27.7 |
| Phek | 931 | 154 | 25.9 | 33.8 |
| Tuensang | 987 | 142 | 20.4 | 24.3 |
| Wokha | 789 | 124 | 24.3 | 35.7 |
| Zunheboto | 778 | 103 | 33.2 | 41.2 |
| Mean |  |  | 22.7 | 30.1 |
| Median |  |  | 22.6 | 30.0 |
| Maximum |  |  | 33.2 | 41.2 |
| Minimum |  |  | 15.3 | 20.8 |
| Interquartile Interval |  |  | 4.95 | 8.71 |

Supplementary Table 22: Prevalence of hypertension in various districts of NCT of Delhi

| States | Sample size | | Age-standardized hypertension  prevalence (%) | |
| --- | --- | --- | --- | --- |
|  | Women | Men | Women | Men |
| **NCT of Delhi** |  |  |  |  |
| Central | 897 | 101 | 26.5 | 41.5 |
| East | 1055 | 158 | 25.8 | 33.8 |
| New Delhi | 924 | 170 | 25.8 | 33.8 |
| North | 929 | 120 | 21.3 | 32.9 |
| North East | 1057 | 155 | 27.2 | 37.0 |
| North West | 996 | 164 | 24.1 | 29.9 |
| Shahdara | 1151 | 174 | 26.1 | 35.0 |
| South | 990 | 147 | 22.3 | 30.4 |
| South East | 1096 | 193 | 23.7 | 27.8 |
| South West | 1064 | 151 | 19.5 | 26.5 |
| West | 1000 | 167 | 23.8 | 34.1 |
| Mean |  |  | 24.2 | 33.0 |
| Median |  |  | 24.1 | 33.8 |
| Maximum |  |  | 27.2 | 41.5 |
| Minimum |  |  | 19.5 | 26.5 |
| Interquartile Interval |  |  | 2.93 | 4.38 |

Supplementary Table 23 : Prevalence of hypertension in various districts of Orissa

| States | Sample size | | Age-standardized hypertension  prevalence (%) | |
| --- | --- | --- | --- | --- |
|  | Women | Men | Women | Men |
| **Orissa** |  |  |  |  |
| Anugul | 964 | 148 | 18.9 | 21.1 |
| Balangir | 989 | 143 | 20.1 | 24.4 |
| Baleshwar | 856 | 100 | 22.9 | 29.5 |
| Bargarh | 891 | 145 | 22.6 | 24.3 |
| Baudh | 970 | 140 | 20.9 | 24.0 |
| Bhadrak | 936 | 106 | 21.7 | 26.1 |
| Cuttack | 845 | 130 | 22.7 | 27.1 |
| Debagarh | 859 | 114 | 20.1 | 27.5 |
| Dhenkanal | 857 | 98 | 25.3 | 26.0 |
| Gajapati | 883 | 109 | 19.7 | 24.9 |
| Ganjam | 955 | 111 | 23.4 | 29.7 |
| Jagatsinghapur | 889 | 103 | 26.2 | 30.2 |
| Jajapur | 959 | 140 | 23.8 | 28.0 |
| Jharsuguda | 904 | 133 | 20.4 | 25.3 |
| Kalahandi | 1034 | 161 | 26.4 | 26.2 |
| Kandhamal | 1015 | 152 | 24.4 | 24.8 |
| Kendrapara | 860 | 111 | 21.9 | 24.8 |
| Kendujhar | 971 | 128 | 24.1 | 25.1 |
| Khordha | 927 | 116 | 20.2 | 25.1 |
| Koraput | 982 | 140 | 21.1 | 24.5 |
| Malkangiri | 1080 | 137 | 17.1 | 20.7 |
| Mayurbhanj | 921 | 123 | 26.1 | 28.8 |
| Nabarangapur | 1017 | 155 | 20.8 | 22.6 |
| Nayagarh | 917 | 132 | 22.0 | 18.8 |
| Nuapada | 944 | 139 | 21.4 | 23.6 |
| Puri | 930 | 120 | 20.8 | 23.3 |
| Rayagada | 910 | 135 | 19.5 | 25.1 |
| Sambalpur | 876 | 133 | 22.2 | 24.5 |
| Subarnapur | 947 | 144 | 22.4 | 20.5 |
| Sundargarh | 883 | 119 | 21.1 | 25.3 |
| Mean |  |  | 22.0 | 25.1 |
| Median |  |  | 21.8 | 25.0 |
| Maximum |  |  | 26.4 | 30.2 |
| Minimum |  |  | 17.1 | 18.8 |
| Interquartile Interval |  |  | 2.79 | 2.05 |

Supplementary Table 24 : Prevalence of hypertension in various districts of Punjab

| States | Sample size | | Age-standardized hypertension  prevalence (%) | |
| --- | --- | --- | --- | --- |
|  | Women | Men | Women | Men |
| **Punjab** |  |  |  |  |
| Amritsar | 965 | 169 | 35.9 | 43.0 |
| Barnala | 956 | 158 | 32.8 | 40.0 |
| Bathinda | 908 | 155 | 32.2 | 45.1 |
| Faridkot | 968 | 142 | 26.1 | 35.4 |
| Fatehgarh Sahib | 905 | 147 | 31.8 | 35.1 |
| Fazilka | 1131 | 187 | 27.9 | 36.2 |
| Firozpur | 1176 | 172 | 29.7 | 33.9 |
| Gurdaspur | 960 | 137 | 36.8 | 39.2 |
| Hoshiarpur | 1027 | 155 | 35.9 | 43.0 |
| Jalandhar | 898 | 141 | 32.5 | 37.5 |
| Kapurthala | 968 | 154 | 32.3 | 36.2 |
| Ludhiana | 817 | 134 | 26.3 | 32.2 |
| Mansa | 1080 | 153 | 30.2 | 42.2 |
| Moga | 1032 | 187 | 31.2 | 35.3 |
| Muktsar | 1022 | 173 | 24.4 | 32.0 |
| Pathankot | 935 | 125 | 33.0 | 32.5 |
| Patiala | 981 | 142 | 28.7 | 34.3 |
| Rupnagar | 969 | 109 | 27.2 | 31.9 |
| Sahibzada Ajit Singh Nagar | 873 | 107 | 27.7 | 34.0 |
| Sangrur | 1052 | 150 | 32.9 | 41.8 |
| Shahid Bhagat Singh Nagar | 1103 | 162 | 32.7 | 42.8 |
| Tarn Taran | 1045 | 137 | 31.1 | 36.1 |
| Mean |  |  | 30.9 | 37.3 |
| Median |  |  | 31.5 | 36.2 |
| Maximum |  |  | 36.8 | 45.1 |
| Minimum |  |  | 24.4 | 31.9 |
| Interquartile Interval |  |  | 4.71 | 7.23 |

Supplementary Table 25 : Prevalence of hypertension in various districts of Pondicherry

| States | Sample size | | Age-standardized hypertension  prevalence (%) | |
| --- | --- | --- | --- | --- |
|  | Women | Men | Women | Men |
| **Pondicherry** |  |  |  |  |
| Karaikal | 890 | 113 | 20.5 | 24.4 |
| Mahe | 981 | 154 | 33.3 | 37.7 |
| Puducherry | 830 | 110 | 22.7 | 31.1 |
| Yanam | 968 | 157 | 32.2 | 33.3 |
| Mean |  |  | 27.2 | 31.6 |
| Median |  |  | 27.5 | 32.2 |
| Maximum |  |  | 33.3 | 37.7 |
| Minimum |  |  | 20.5 | 24.4 |
| Interquartile Interval |  |  | 10.36 | 5.00 |

Supplementary Table 26 : Prevalence of hypertension in various districts of Rajasthan

| States | Sample size | | Age-standardized hypertension  prevalence (%) | |
| --- | --- | --- | --- | --- |
|  | Women | Men | Women | Men |
| **Rajasthan** |  |  |  |  |
| Ajmer | 1082 | 146 | 14.5 | 13.1 |
| Alwar | 1182 | 152 | 20.3 | 25.8 |
| Banswara | 1172 | 170 | 17.1 | 19.4 |
| Baran | 1301 | 227 | 12.6 | 15.6 |
| Barmer | 1621 | 237 | 8.5 | 10.9 |
| Bharatpur | 1225 | 168 | 19.1 | 23.6 |
| Bhilwara | 1147 | 171 | 15.4 | 15.7 |
| Bikaner | 1473 | 241 | 12.0 | 11.8 |
| Bundi | 1253 | 190 | 17.2 | 19.0 |
| Chittaurgarh | 1090 | 141 | 18.3 | 21.1 |
| Churu | 1493 | 225 | 21.2 | 25.4 |
| Dausa | 1332 | 208 | 12.2 | 16.1 |
| Dhaulpur | 1220 | 187 | 13.4 | 15.1 |
| Dungarpur | 1359 | 223 | 12.9 | 14.9 |
| Ganganagar | 1409 | 223 | 16.2 | 18.3 |
| Hanumangarh | 1372 | 223 | 20.0 | 22.6 |
| Jaipur | 1241 | 198 | 14.7 | 16.5 |
| Jaisalmer | 1537 | 237 | 9.7 | 15.8 |
| Jalor | 1318 | 193 | 10.4 | 11.5 |
| Jhalawar | 1209 | 177 | 15.0 | 17.3 |
| Jhunjhunun | 1388 | 186 | 20.7 | 25.6 |
| Jodhpur | 1535 | 239 | 13.1 | 16.5 |
| Karauli | 1167 | 176 | 10.6 | 13.2 |
| Kota | 1278 | 158 | 15.9 | 20.2 |
| Nagaur | 1601 | 221 | 15.6 | 17.5 |
| Pali | 1236 | 159 | 13.0 | 16.5 |
| Pratapgarh | 1199 | 177 | 16.5 | 18.7 |
| Rajsamand | 1153 | 152 | 15.1 | 15.4 |
| Sawai Madhopur | 1118 | 177 | 15.4 | 19.8 |
| Sikar | 1458 | 203 | 19.4 | 22.0 |
| Sirohi | 1304 | 190 | 14.2 | 18.7 |
| Tonk | 1240 | 187 | 16.7 | 20.1 |
| Udaipur | 1277 | 191 | 14.4 | 16.5 |
| Mean |  |  | 15.2 | 17.9 |
| Median |  |  | 15.1 | 17.3 |
| Maximum |  |  | 21.2 | 25.8 |
| Minimum |  |  | 8.5 | 10.9 |
| Interquartile Interval |  |  | 4.10 | 4.48 |

Supplmentary Table 27: Prevalence of hypertension in various districts of Sikkim

| States | Sample size | | Age-standardized hypertension | |
| --- | --- | --- | --- | --- |
|  |  |  | prevalence (%) | |
|  | Women | Men | Women | Men |
| **Sikkim** |  |  |  |  |
| East District | 724 | 93 | 31.6 | 34.2 |
| North District | 768 | 112 | 39.0 | 47.7 |
| South District | 745 | 114 | 41.0 | 49.6 |
| West District | 1034 | 150 | 32.7 | 45.1 |
| Mean |  |  | 36.1 | 44.1 |
| Median |  |  | 35.8 | 46.4 |
| Maximum |  |  | 41.0 | 49.6 |
| Minimum |  |  | 31.6 | 34.2 |
| Interquartile Interval |  |  | 7.06 | 5.84 |

Supplementary Table 28: Prevalence of hypertension in various districts of Tamilnadu

| States | Sample size | | Age-standardized hypertension | |
| --- | --- | --- | --- | --- |
|  |  |  | prevalence (%) | |
|  | Women | Men | Women | Men |
| **Tamilnadu** |  |  |  |  |
| Ariyalur | 765 | 93 | 20.6 | 27.5 |
| Chennai | 660 | 101 | 28.8 | 33.2 |
| Coimbatore | 763 | 95 | 29.2 | 33.3 |
| Cuddalore | 859 | 98 | 20.1 | 27.0 |
| Dharmapuri | 827 | 101 | 25.8 | 30.8 |
| Dindigul | 951 | 142 | 25.7 | 34.2 |
| Erode | 774 | 110 | 29.5 | 36.1 |
| Kancheepuram | 796 | 102 | 24.3 | 32.0 |
| Kanniyakumari | 711 | 103 | 24.4 | 30.2 |
| Karur | 689 | 93 | 24.1 | 29.1 |
| Krishnagiri | 909 | 130 | 23.5 | 25.3 |
| Madurai | 844 | 131 | 25.6 | 31.8 |
| Nagapattinam | 794 | 96 | 24.5 | 32.1 |
| Namakkal | 700 | 109 | 25.6 | 31.5 |
| Perambalur | 724 | 87 | 19.8 | 26.1 |
| Pudukkottai | 871 | 101 | 23.0 | 30.2 |
| Ramanathapuram | 913 | 120 | 23.3 | 28.2 |
| Salem | 754 | 99 | 26.6 | 34.0 |
| Sivaganga | 887 | 106 | 22.0 | 26.0 |
| Thanjavur | 687 | 83 | 28.8 | 34.6 |
| The Nilgiris | 850 | 120 | 34.2 | 32.9 |
| Theni | 979 | 139 | 28.7 | 35.6 |
| Thiruvallur | 775 | 124 | 26.4 | 32.4 |
| Thiruvarur | 783 | 98 | 23.8 | 31.0 |
| Thoothukkudi | 774 | 99 | 24.5 | 28.2 |
| Tiruchirappalli | 764 | 89 | 21.4 | 28.9 |
| Tirunelveli | 777 | 92 | 22.1 | 25.5 |
| Tiruppur | 726 | 85 | 26.8 | 28.9 |
| Tiruvannamalai | 786 | 108 | 19.2 | 25.8 |
| Vellore | 886 | 108 | 25.9 | 25.6 |
| Viluppuram | 812 | 109 | 19.0 | 26.1 |
| Virudhunagar | 860 | 101 | 20.2 | 22.9 |
| Mean |  |  | 24.6 | 29.9 |
| Median |  |  | 24.4 | 30.2 |
| Maximum |  |  | 34.2 | 36.1 |
| Minimum |  |  | 19.0 | 22.9 |
| Interquartile Interval |  |  | 4.40 | 5.80 |

Supplementary Table 29 : Prevalence of hypertension in various districts of Telangana

| States | Sample size | | Age-standardized hypertension | |
| --- | --- | --- | --- | --- |
|  |  |  | prevalence (%) | |
|  | Women | Men | Women | Men |
| **Telangana** |  |  |  |  |
| Adilabad | 965 | 156 | 23.1 | 26.4 |
| Bhadradri Kothagudem | 945 | 136 | 29.6 | 30.1 |
| Hyderabad | 642 | 84 | 30.2 | 41.7 |
| Jagitial | 885 | 99 | 25.5 | 31.3 |
| Jangoan | 843 | 108 | 26.1 | 29.0 |
| Jayashankar Bhupalapally | 852 | 124 | 28.4 | 31.5 |
| Jogulamba Gadwal | 975 | 125 | 21.5 | 29.7 |
| Kamareddy | 857 | 134 | 27.7 | 29.1 |
| Karimnagar | 853 | 114 | 25.0 | 34.7 |
| Khammam | 906 | 132 | 27.6 | 30.8 |
| Komaram Bheem Asifabad | 934 | 136 | 21.8 | 27.2 |
| Mahabubabad | 903 | 121 | 21.9 | 28.8 |
| Mahabubnagar | 1006 | 137 | 24.2 | 26.5 |
| Mancherial | 846 | 118 | 27.9 | 32.0 |
| Medak | 883 | 128 | 26.7 | 28.9 |
| MedchalMalkajgiri | 825 | 107 | 26.5 | 34.8 |
| Nagarkurnool | 915 | 136 | 24.6 | 27.3 |
| Nalgonda | 879 | 129 | 19.6 | 25.7 |
| Nirmal | 851 | 107 | 25.1 | 28.5 |
| Nizamabad | 891 | 107 | 27.1 | 31.2 |
| Peddapalli | 861 | 142 | 27.7 | 34.7 |
| Rajanna Sircilla | 901 | 125 | 30.2 | 36.0 |
| Ranga Reddy | 935 | 104 | 28.5 | 35.0 |
| Sangareddy | 911 | 119 | 25.7 | 32.6 |
| Siddipet | 832 | 116 | 26.6 | 36.9 |
| Suryapet | 851 | 125 | 25.2 | 27.3 |
| Vikarabad | 929 | 132 | 27.0 | 30.3 |
| Wanaparthy | 965 | 146 | 22.5 | 25.5 |
| Warangal Rural | 830 | 123 | 26.3 | 29.4 |
| Warangal Urban | 900 | 138 | 24.5 | 32.7 |
| Yadadri Bhuvanagiri | 947 | 155 | 24.8 | 32.3 |
| Mean |  |  | 25.8 | 30.9 |
| Median |  |  | 26.1 | 30.3 |
| Maximum |  |  | 30.2 | 41.7 |
| Minimum |  |  | 19.6 | 25.5 |
| Interquartile Interval |  |  | 3.14 | 3.97 |

Supplementary Table 30: Prevalence of hypertension in various districts of Tripura

| States | Sample size | | Age-standardized hypertension | |
| --- | --- | --- | --- | --- |
|  |  |  | prevalence (%) | |
|  | Women | Men | Women | Men |
| **Tripura** |  |  |  |  |
| Dhalai | 979 | 130 | 14.5 | 17.4 |
| Gomati | 851 | 109 | 19.8 | 21.4 |
| Khowai | 872 | 137 | 19.0 | 21.4 |
| North Tripura | 1039 | 125 | 18.8 | 19.3 |
| Sepahijala | 892 | 121 | 19.3 | 19.9 |
| South Tripura | 866 | 96 | 22.4 | 24.2 |
| Unakoti | 950 | 121 | 21.0 | 22.1 |
| West Tripura | 865 | 151 | 25.1 | 27.1 |
| Mean |  |  | 20.0 | 21.6 |
| Median |  |  | 19.5 | 21.4 |
| Maximum |  |  | 25.1 | 27.1 |
| Minimum |  |  | 14.5 | 17.4 |
| Interquartile Interval |  |  | 2.39 | 2.83 |

Supplementary Table 31 : Prevalence of hypertension in various districts of Uttrakhand

| States | Sample size | | Age-standardized hypertension | |
| --- | --- | --- | --- | --- |
|  |  |  | prevalence (%) | |
|  | Women | Men | Women | Men |
| **Uttrakhand** |  |  |  |  |
| Almora | 938 | 91 | 24.6 | 33.2 |
| Bageshwar | 1008 | 113 | 20.2 | 34.9 |
| Chamoli | 1016 | 114 | 19.7 | 28.5 |
| Champawat | 1177 | 147 | 19.9 | 29.7 |
| Dehradun | 979 | 93 | 23.7 | 31.9 |
| Garhwal | 836 | 80 | 22.1 | 34.3 |
| Hardwar | 968 | 121 | 26.2 | 33.9 |
| Nainital | 1078 | 155 | 24.0 | 30.4 |
| Pithoragarh | 1085 | 145 | 23.3 | 32.9 |
| Rudraprayag | 1032 | 108 | 18.7 | 31.7 |
| Tehri Garhwal | 971 | 111 | 20.0 | 34.0 |
| Udham Singh Nagar | 1143 | 143 | 22.2 | 30.2 |
| Uttarkashi | 1049 | 165 | 19.0 | 24.4 |
| Mean |  |  | 21.8 | 31.5 |
| Median |  |  | 22.1 | 31.9 |
| Maximum |  |  | 26.2 | 34.9 |
| Minimum |  |  | 18.7 | 24.4 |
| Interquartile Interval |  |  | 3.78 | 3.67 |

Supplementary Table 32 : Prevalence of hypertension in various districts of Uttar Pradesh

|  | Sample size | | Age-standardized hypertension | |
| --- | --- | --- | --- | --- |
|  |  |  | prevalence (%) | |
|  | Women | Men | Women | Men |
| **Uttar Pradesh** |  |  |  |  |
| Agra | 1226 | 159 | 16.8 | 20.9 |
| Aligarh | 1073 | 145 | 18.2 | 22.0 |
| Allahabad | 1124 | 123 | 12.6 | 17.7 |
| Ambedkar Nagar | 1259 | 130 | 17.2 | 22.0 |
| Amethi | 1255 | 124 | 19.2 | 21.2 |
| Auraiya | 1309 | 166 | 20.0 | 22.1 |
| Azamgarh | 1354 | 146 | 19.4 | 28.7 |
| Baghpat | 1304 | 203 | 21.4 | 25.5 |
| Bahraich | 1128 | 129 | 24.2 | 28.7 |
| Ballia | 1254 | 144 | 17.1 | 20.7 |
| Balrampur | 1235 | 141 | 23.2 | 25.3 |
| Banda | 1189 | 203 | 20.1 | 18.9 |
| Bara Banki | 1186 | 184 | 19.1 | 22.7 |
| Bareilly | 1088 | 99 | 16.4 | 17.6 |
| Basti | 1362 | 147 | 19.2 | 25.0 |
| Bijnor | 1348 | 178 | 19.7 | 24.3 |
| Budaun | 1134 | 166 | 13.0 | 13.8 |
| Bulandshahr | 1105 | 140 | 21.2 | 26.2 |
| Chandauli | 1272 | 141 | 18.9 | 23.6 |
| Chitrakoot | 1176 | 173 | 15.1 | 16.4 |
| Deoria | 1317 | 157 | 17.6 | 21.0 |
| Etah | 1267 | 179 | 16.4 | 19.2 |
| Etawah | 1225 | 183 | 16.7 | 19.7 |
| Faizabad | 1213 | 137 | 14.9 | 17.7 |
| Farrukhabad | 1276 | 175 | 13.8 | 14.8 |
| Fatehpur | 1206 | 175 | 17.0 | 18.0 |
| Firozabad | 1178 | 181 | 18.1 | 20.5 |
| Gautam Buddha Nagar | 863 | 95 | 16.4 | 20.9 |
| Ghaziabad | 805 | 76 | 19.3 | 24.5 |
| Ghazipur | 1363 | 203 | 14.2 | 20.3 |
| Gonda | 1292 | 152 | 22.9 | 28.6 |
| Gorakhpur | 1360 | 135 | 16.8 | 20.7 |
| Hamirpur | 1389 | 206 | 16.7 | 18.8 |
| Hapur | 1357 | 222 | 22.9 | 25.7 |
| Hardoi | 1206 | 188 | 18.1 | 19.9 |
| Jalaun | 1373 | 198 | 16.8 | 20.0 |
| Jaunpur | 1561 | 176 | 16.0 | 19.7 |
| Jhansi | 1326 | 229 | 15.7 | 17.3 |
| Jyotiba Phule Nagar | 1411 | 165 | 19.1 | 24.6 |
| Kannauj | 1317 | 198 | 14.9 | 14.4 |
| Kanpur Dehat | 1213 | 144 | 12.6 | 16.5 |
| Kanpur Nagar | 1150 | 177 | 17.3 | 16.2 |
| Kanshiram Nagar | 1143 | 163 | 16.5 | 19.5 |
| Kaushambi | 1100 | 140 | 9.4 | 10.0 |
| Kheri | 1280 | 169 | 20.1 | 22.4 |
| Kushinagar | 1312 | 138 | 16.4 | 21.9 |
| Lalitpur | 1484 | 230 | 17.2 | 17.9 |
| Lucknow | 958 | 117 | 21.2 | 22.8 |
| Mahamaya Nagar | 1234 | 146 | 22.4 | 26.4 |
| Mahoba | 1347 | 226 | 15.0 | 15.7 |
| Mahrajganj | 1316 | 120 | 19.6 | 24.9 |
| Mainpuri | 1258 | 168 | 14.4 | 17.9 |
| Mathura | 1175 | 162 | 21.1 | 25.4 |
| Mau | 1310 | 189 | 23.9 | 29.9 |
| Meerut | 1212 | 169 | 20.0 | 26.2 |
| Mirzapur | 1176 | 147 | 21.7 | 29.0 |
| Moradabad | 1264 | 160 | 21.3 | 25.6 |
| Muzaffarnagar | 1201 | 179 | 21.6 | 24.2 |
| Pilibhit | 1040 | 142 | 19.5 | 17.5 |
| Pratapgarh | 1267 | 96 | 17.6 | 22.3 |
| Rae Bareli | 1258 | 148 | 19.4 | 17.9 |
| Rampur | 1092 | 145 | 17.7 | 21.0 |
| Saharanpur | 1318 | 183 | 23.0 | 24.3 |
| Sambhal | 1314 | 199 | 19.3 | 22.7 |
| Sant Kabir Nagar | 1266 | 120 | 22.5 | 28.2 |
| Sant Ravidas Nagar (Bhadohi) | 1352 | 118 | 14.9 | 23.0 |
| Shahjahanpur | 1286 | 220 | 16.1 | 14.5 |
| Shamli | 1300 | 180 | 21.2 | 28.6 |
| Shrawasti | 1233 | 154 | 18.4 | 18.7 |
| Siddharthnagar | 1396 | 127 | 19.1 | 23.5 |
| Sitapur | 1212 | 148 | 17.8 | 23.0 |
| Sonbhadra | 1158 | 168 | 19.0 | 21.4 |
| Sultanpur | 1282 | 133 | 20.6 | 24.3 |
| Unnao | 1128 | 152 | 22.0 | 21.4 |
| Varanasi | 1403 | 195 | 19.0 | 27.1 |
| Mean |  |  | 18.3 | 21.6 |
| Median |  |  | 18.4 | 21.4 |
| Maximum |  |  | 24.2 | 29.9 |
| Minimum |  |  | 9.4 | 10.0 |
| Interquartile Interval |  |  | 3.66 | 5.81 |

Supplementary Table 33: Prevalence of hypertension in various districts of West Bengal

| States | Sample size | | Age-standardized hypertension | |
| --- | --- | --- | --- | --- |
|  |  |  | prevalence (%) | |
|  | Women | Men | Women | Men |
| **West Bengal** |  |  |  |  |
| Bankura | 997 | 133 | 21.0 | 20.3 |
| Birbhum | 1161 | 168 | 17.5 | 14.1 |
| Dakshin Dinajpur | 1137 | 163 | 17.6 | 16.2 |
| Darjiling | 1058 | 163 | 29.2 | 29.5 |
| Haora | 1067 | 153 | 24.0 | 26.8 |
| Hugli | 1020 | 136 | 24.1 | 26.7 |
| Jalpaiguri | 1101 | 149 | 21.2 | 23.7 |
| Koch Bihar | 1095 | 157 | 21.7 | 20.8 |
| Kolkata | 921 | 138 | 23.9 | 23.8 |
| Maldah | 1113 | 150 | 19.3 | 17.8 |
| Murshidabad | 1144 | 159 | 18.7 | 16.3 |
| Nadia | 1034 | 146 | 21.2 | 16.0 |
| North Twenty Four Pargana | 1055 | 148 | 19.0 | 17.1 |
| Paschim Bardhaman | 1190 | 184 | 17.6 | 20.7 |
| Paschim Medinipur | 1002 | 137 | 19.3 | 19.0 |
| Purba Bardhaman | 1088 | 163 | 21.2 | 18.4 |
| Purba Medinipur | 957 | 131 | 18.1 | 21.3 |
| Puruliya | 1050 | 138 | 17.5 | 20.4 |
| South Twenty Four Pargana | 1089 | 148 | 21.5 | 19.1 |
| Uttar Dinajpur | 1129 | 157 | 16.3 | 23.8 |
| Mean |  |  | 20.5 | 20.6 |
| Median |  |  | 20.2 | 20.4 |
| Maximum |  |  | 29.2 | 29.5 |
| Minimum |  |  | 16.3 | 14.1 |
| Interquartile Interval |  |  | 3.57 | 6.09 |

Supplementary Table 34: Prevalence of hypertension in various districts of Union Territory

| **States** | **Sample size** | | **Age-standardized hypertension** | |
| --- | --- | --- | --- | --- |
|  |  |  | **prevalence (%)** | |
|  | **Women** | **Men** | **Women** | **Men** |
| Andaman & Nicobar Island |  |  |  |  |
| North & Middle Andaman | 789 | 108 | 27.4 | 32.2 |
| South Andaman | 844 | 134 | 23.0 | 26.9 |
| Nicobars | 764 | 125 | 35.4 | 47.0 |
| Mean |  |  | 28.6 | 35.4 |
| Median |  |  | 27.4 | 32.2 |
| Maximum |  |  | 35.4 | 47.0 |
| Minimum |  |  | 23.0 | 26.9 |
| Interquartile Interval |  |  | 6.21 | 10.04 |
| Dadra & Nagar Haveli | 1002 | 174 | 13.3 | 13.5 |
| Daman | 733 | 152 | 18.4 | 19.9 |
| Diu | 978 | 101 | 18.1 | 17.7 |
| Ladakh (Kargil) |  |  | 18.7 | 20.1 |
| Ladakh (Leh) |  |  | 12.8 | 14.7 |
| Lakshadweep | 1234 | 135 | 24.8 | 24.7 |

**Supplementary Table 35: Correlation matrix of various factors**

| Factors | Hypertension (%) Women | MPI | Women  Literacy | Electricity Access | Improved Drinking Water | Improved Sanitation | Health Insurance | Clean Fuel | Tobacco Use Women | Skilled Delivery | Modern Contraception Use | Fully Vaccinated | Child Marriage | Teenage Pregnancy | Anaemia (Pregnancy) | Anaemia  Women | Childhood Stunting | Childhood wasted | Underweight (under five) | Overweight (under five) | Overweight  Adult | hyperglycaemia |
| --- | --- | --- | --- | --- | --- | --- | --- | --- | --- | --- | --- | --- | --- | --- | --- | --- | --- | --- | --- | --- | --- | --- |
| Hypertension (%)  Women | 1.00 | -.497** | .406** | .281** | .093* | .472** | 0.00 | .444** | -.116** | .275** | .224** | .178** | -.342** | -.147** | -.169** | -.276** | -.422** | -.228** | -.367** | .107** | .567** | .417** |
| MPI | -.497** | 1.00 | -.741** | -.578** | -.257** | -.712** | -.078* | -.751** | .258** | -.585** | -.341** | -.278** | .563** | .342** | .348** | .287** | .669** | .288** | .610** | -.259** | -.723** | -.428** |
| Women Literacy | .406** | -.741** | 1.00 | .336** | -0.03 | .666** | 0.01 | .449** | 0.03 | .292** | .081* | .177** | -.607** | -.282** | -.377** | -.365** | -.543** | -.286** | -.560** | .228** | .541** | .435** |
| Electricity Access | .281** | -.578** | .336** | 1.00 | .132** | .323** | .129** | .396** | -.250** | .396** | .383** | .304** | -.236** | -.139** | -0.05 | -.086* | -.429** | -.120** | -.293** | 0.06 | .389** | .280** |
| Improved drinking  water | .093* | -.257** | -0.03 | .132** | 1.00 | .113** | -.101** | .337** | -.344** | .289** | .207** | .124** | -0.05 | -.110** | 0.06 | 0.03 | -0.04 | -.103** | -0.04 | 0.00 | .329** | .154** |
| Improved sanitation | .472** | -.712** | .666** | .323** | .113** | 1.00 | .080* | .463** | -0.07 | .201** | .151** | .103** | -.571** | -.351** | -.450** | -.450** | -.503** | -.340** | -.583** | .256** | .481** | .245** |
| Health Insurance | 0.00 | -.078* | 0.01 | .129** | -.101** | .080* | 1.00 | -0.07 | .096* | .193** | .253** | .203** | 0.01 | 0.03 | 0.05 | .079* | -.120** | -0.04 | -0.05 | -.082* | -.148** | .089* |
| Clean Fuel | .444** | -.751** | .449** | .396** | .337** | .463** | -0.07 | 1.00 | -.263** | .497** | .305** | .155** | -.359** | -.230** | -.243** | -.230** | -.413** | -.126** | -.343** | .181** | .733** | .440** |
| Tobacco Use Women | -.116** | .258** | 0.03 | -.250** | -.344** | -0.07 | .096* | -.263** | 1.00 | -.251** | -.347** | -.128** | .101** | .253** | -0.02 | -0.05 | .097* | -0.02 | -0.01 | .119** | -.292** | -.129** |
| Skilled Delivery | .275** | -.585** | .292** | .396** | .289** | .201** | .193** | .497** | -.251** | 1.00 | .456** | .447** | -.200** | -.196** | 0.04 | .102** | -.398** | -0.03 | -.186** | 0.03 | .449** | .381** |
| Modern Contraception  Use | .224** | -.341** | .081* | .383** | .207** | .151** | .253** | .305** | -.347** | .456** | 1.00 | .436** | -0.07 | -.078* | 0.00 | 0.00 | -.170** | .133** | 0.04 | -0.02 | .140** | .101** |
| Fully Vaccinated | .178** | -.278** | .177** | .304** | .124** | .103** | .203** | .155** | -.128** | .447** | .436** | 1.00 | -.207** | -.147** | 0.08 | .113** | -.195** | 0.01 | -0.06 | 0.02 | .176** | .155** |
| Child Marriage | -.342** | .563** | -.607** | -.236** | -0.05 | -.571** | 0.01 | -.359** | .101** | -.200** | -0.07 | -.207** | 1.00 | .738** | .397** | .361** | .445** | .296** | .499** | -.252** | -.412** | -.100** |
| Teenage Pregnancy | -.147** | .342** | -.282** | -.139** | -.110** | -.351** | 0.03 | -.230** | .253** | -.196** | -.078* | -.147** | .738** | 1.00 | .351** | .316** | .241** | .153** | .275** | -.102** | -.188** | .128** |
| Anaemia Pregnant | -.169** | .348** | -.377** | -0.05 | 0.06 | -.450** | 0.05 | -.243** | -0.02 | 0.04 | 0.00 | 0.08 | .397** | .351** | 1.00 | .745** | .315** | .202** | .411** | -.259** | -.132** | .150** |
| Anaemia Woman | -.276** | .287** | -.365** | -.086* | 0.03 | -.450** | .079* | -.230** | -0.05 | .102** | 0.00 | .113** | .361** | .316** | .745** | 1.00 | .239** | .303** | .411** | -.102** | -.223** | 0.00 |
| Childhood Stunting | -.422** | .669** | -.543** | -.429** | -0.04 | -.503** | -.120** | -.413** | .097* | -.398** | -.170** | -.195** | .445** | .241** | .315** | .239** | 1.00 | .240** | .702** | -.149** | -.561** | -.367** |
| Childhood Wasted | -.228** | .288** | -.286** | -.120** | -.103** | -.340** | -0.04 | -.126** | -0.02 | -0.03 | .133** | 0.01 | .296** | .153** | .202** | .303** | .240** | 1.00 | .719** | -.156** | -.442** | -.112** |
| Underweight  (Under Five) | -.367** | .610** | -.560** | -.293** | -0.04 | -.583** | -0.05 | -.343** | -0.01 | -.186** | 0.04 | -0.06 | .499** | .275** | .411** | .411** | .702** | .719** | 1.00 | -.396** | -.580** | -.188** |
| Overweight  (Under Five) | .107** | -.259** | .228** | 0.06 | 0.00 | .256** | -.082* | .181** | .119** | 0.03 | -0.02 | 0.02 | -.252** | -.102** | -.259** | -.102** | -.149** | -.156** | -.396** | 1.00 | .143** | -.100** |
| Overweight Adult | .567** | -.723** | .541** | .389** | .329** | .481** | -.148** | .733** | -.292** | .449** | .140** | .176** | -.412** | -.188** | -.132** | -.223** | -.561** | -.442** | -.580** | .143** | 1.00 | .616** |
| Hyperglycaemia | .417** | -.428** | .435** | .280** | .154** | .245** | .089* | .440** | -.129** | .381** | .101** | .155** | -.100** | .128** | .150** | 0.00 | -.367** | -.112** | -.188** | -.100** | .616** | 1.00 |

**Supplementary Figure 1: Hypertension prevalence (%) (median of district level prevalence) in different states of India**

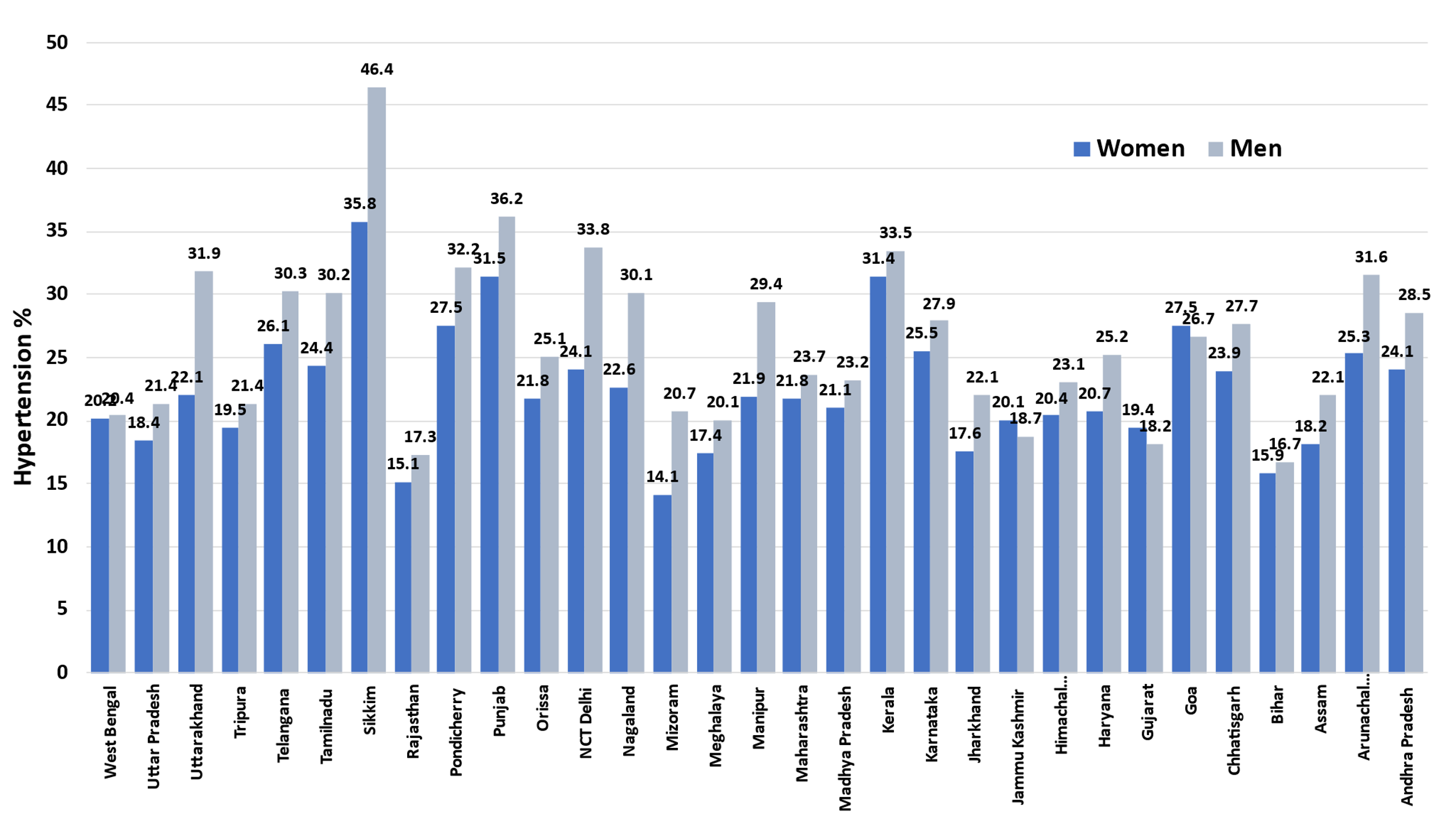

**Supplementary Figure 2: Hypertension prevalence (%) among women and men <30 years in different states of India**

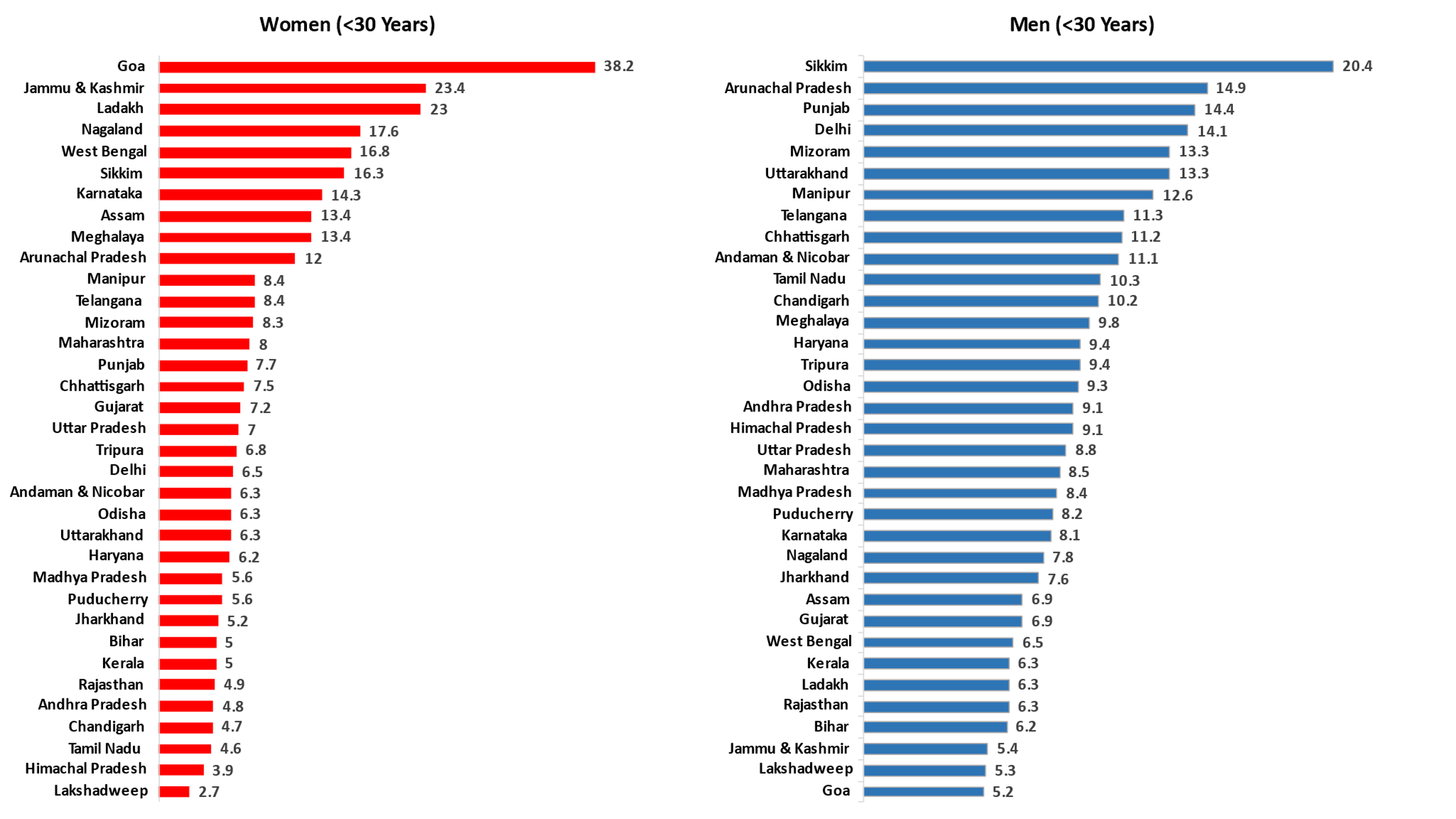

**Supplementary Figure 3 (a-c): Hypertension prevalence (%) among women in districts with low, medium and high levels of (a) socioeconomic determinants, (b) healthcare delivery and female-specific factors, and (c) nutrition, tobacco and clinical variables**

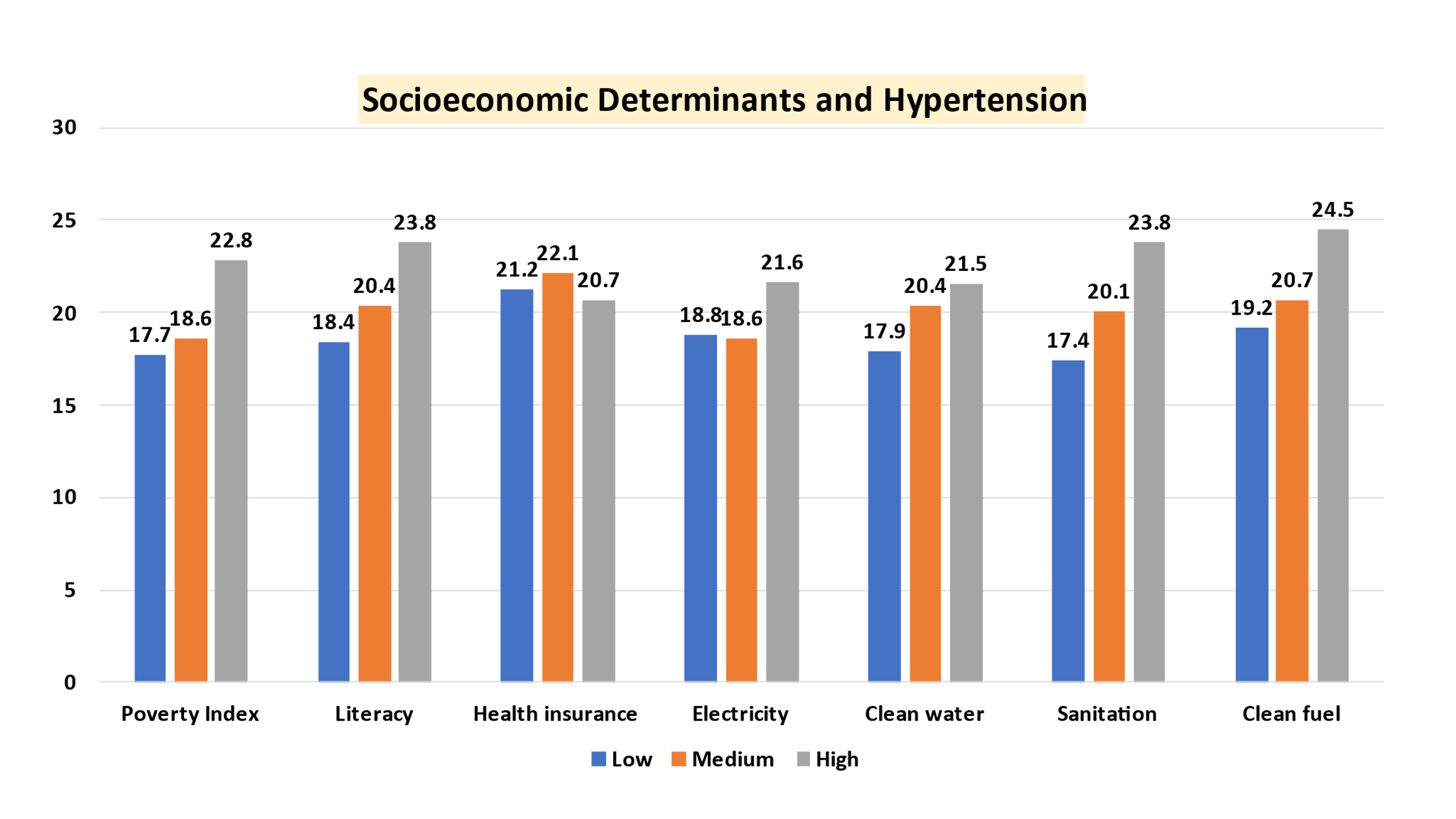

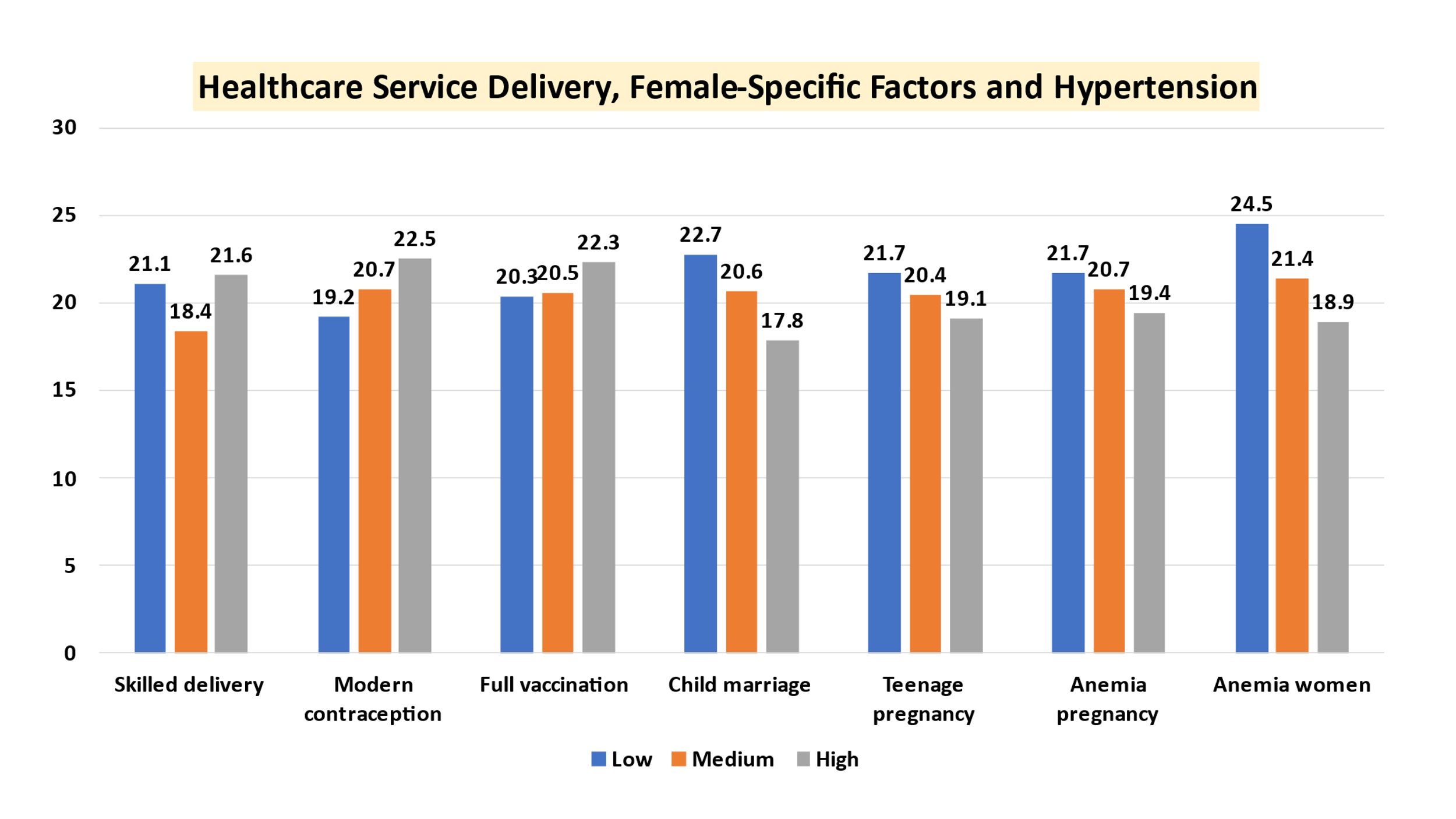

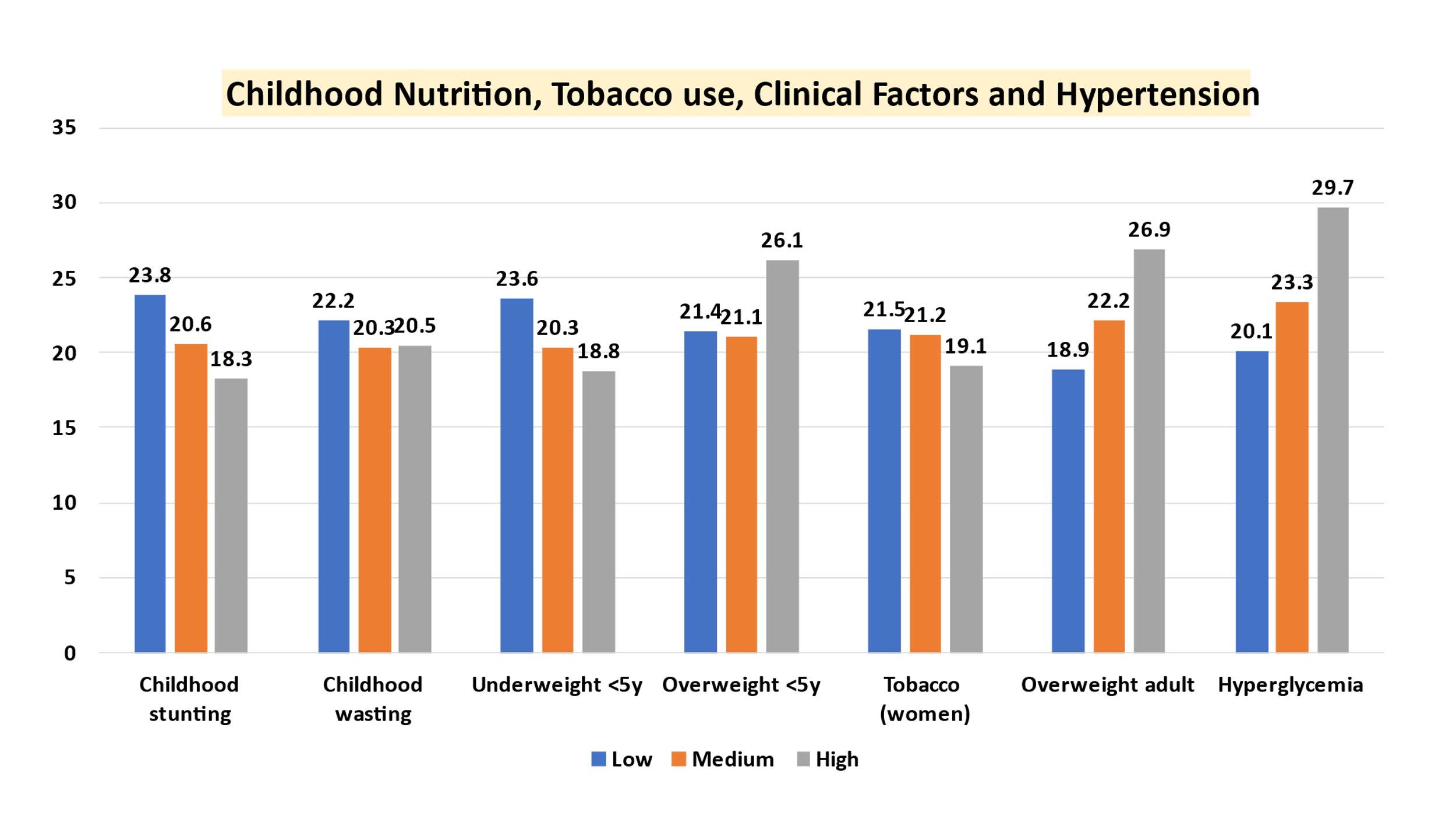
